# CD8^+^ T-cell landscape in Indigenous and non-Indigenous people restricted by influenza mortality-associated HLA-A*24:02 allomorph

**DOI:** 10.1101/2020.10.02.20206086

**Authors:** Luca Hensen, Patricia T. Illing, E. Bridie Clemens, Thi H.O. Nguyen, Marios Koutsakos, Carolien E. van de Sandt, Nicole A. Mifsud, Andrea Nguyen, Christopher Szeto, Brendon Y. Chua, Hanim Halim, Simone Rizzetto, Fabio Luciani, Liyen Loh, Emma J. Grant, Phillipa M. Saunders, Andrew G Brooks, Steve Rockman, Tom C. Kotsimbos, Allen C. Cheng, Michael Richards, Glen P. Westall, Linda M. Wakim, Thomas Loudovaris, Stuart I. Mannering, Michael Elliott, Stuart G. Tangye, David C Jackson, Katie L Flanagan, Jamie Rossjohn, Stephanie Gras, Jane Davies, Adrian Miller, Steven Y.C. Tong, Anthony W. Purcell, Katherine Kedzierska

## Abstract

Indigenous people worldwide are at high-risk of developing severe influenza disease. HLA-A*24:02 allele, highly prevalent in Indigenous populations, is associated with influenza-induced mortality, although the basis for this association is unclear. We defined CD8^+^ T-cell immune landscapes against influenza A (IAV) and B (IBV) viruses in HLA-A*24:02-expressing Indigenous and non-Indigenous individuals, human tissues, influenza-infected patients and HLA-A*24:02-transgenic mice. We identified immunodominant protective CD8^+^ T-cell epitopes, one towards IAV and six towards IBV, with A24/PB2_550-558_-specific CD8^+^ T-cells cells being cross-reactive between IAV and IBV. Memory CD8^+^ T-cells towards these specificities were present in blood (CD27^+^CD45RA^-^ phenotype) and tissues (CD103^+^CD69^+^ phenotype) of healthy subjects, and effector CD27^-^CD45RA^-^PD-1^+^CD38^+^CD8^+^ T-cells in IAV/IBV patients. Our data present the first evidence of influenza-specific CD8^+^ T-cell responses in Indigenous Australians, and advocate for T-cell-mediated vaccines that target and boost the breadth of IAV/IBV-specific CD8^+^ T-cells to protect high-risk HLA-A*24:02-expressing Indigenous and non-Indigenous populations from severe influenza disease.

**One Sentence Summary:** Influenza-specific CD8^+^ T-cell specificities restricted by HLA-A*24:02.

## INTRODUCTION

Newly emerging respiratory viruses pose a major global pandemic threat, leading to significant morbidity and mortality, as exemplified by 2019 SARS-CoV2, avian influenza H5N1 and H7N9 viruses, and the 1918-1919 H1N1 pandemic catastrophe. Influenza A viruses (IAV) can cause sporadic pandemics when a virus reassorts and rapidly spreads across continents, causing millions of infections and deaths^1^. Additionally, seasonal epidemics caused by co-circulating IAV and influenza B viruses (IBV) result in 3-5 million cases of severe disease and 290,000-650,000 deaths annually^2,3^. Severe illness and death from seasonal and pandemic influenza occur disproportionately in high risk individuals, including Indigenous populations. This is most evident when unpredicted seasonal or pandemic viruses emerge in the human circulation. During the 1918-1919 influenza pandemic, 100% of Alaskan adults died in some isolated villages, while only school-aged children survived^4^. Western Samoa was the hardest hit with a total population loss of 19-22% ^5^. As many as 10-20% of Indigenous Australians died from pandemic influenza in 1919^6^ in comparison to <1% of other Australians, with some reports showing up to 50% mortality in Indigenous Australian communities^7^.

During the 2009 A/H1N1 influenza pandemic, Indigenous populations worldwide were more susceptible to influenza-related morbidity and mortality. Hospitalization and morbidity rates were markedly increased in Indigenous Australians^9,10^, with 16% of hospitalised pandemic H1N1 (pH1N1) patients in Australia being Indigenous. The relative risk for Indigenous Australians compared to non-Indigenous Australians for hospitalization, ICU admission or death was 6.6, 6.2, or 5.2 times higher, respectively^11^. This was mirrored in Indigenous populations globally, including American Indians and Alaskan Native people (4-fold higher mortality rate compared to non-Indigenous Americans)^12^, native Brazilians (2-fold higher hospitalization rate)^13^, New Zealand Maori (5-fold higher hospitalization rate) and Pacific Islanders (7-fold higher hospitalization rate)^14,15^. Although the impact of influenza pandemics is more pronounced in Indigenous populations globally, these disproportionate hospitalization rates also occur during seasonal infections. During 2010-2013, Indigenous Australians had increased influenza-related hospitalizations across all age groups (1.2-4.3-fold higher compared to non-Indigenous)^16^. Indigenous populations, especially Australians and Alaskans, are also predicted to be at greater risk from severe disease caused by the avian-derived H7N9 influenza virus, with mortality rates being >30% and hospitalization >99% in China^17^. While higher influenza infection rates could relate to overcrowded living conditions, increased severity and prolonged hospitalization most likely reflect differences in pre-existing immunity that facilitates recovery. However, the underlying immunological and host factors that account for severe influenza disease in Indigenous individuals are far from clear.

Antibody-based vaccines towards variable surface glycoproteins, hemagglutinin (HA) and neuraminidase (NA), are an effective way to combat seasonal infections, yet they fail to provide effective protection when a new, antigenically different IAV emerges^20^. In the absence of antibodies, recall of pre-existing cross-protective memory CD8^+^ T-cells minimizes the effects of a novel IAV, leading to a milder disease after infection with distinct strains^21–26^. Such pre-existing memory CD8^+^ T-cells provide broadly heterotypic or cross-reactive protection and can recognize numerous the IAV, IBV and influenza C viruses capable of infecting humans^27^, promoting rapid host recovery. During the 2013 H7N9 IAV outbreak in China, recovery from severe H7N9 disease was associated with early CD8^+^ T-cell responses^24,28^. Patients discharged early after hospitalization had early (day10) robust H7N9-specific CD8^+^ T-cells responses, while those with prolonged hospital stays showed late (day19) recruitment of CD8^+^ and CD4^+^ T-cells. Thus, with the continuing threat of unpredicted influenza strains, there is a need for targeting cellular immunity that provides effective, long-lasting and cross-strain protective immunity, especially for high risk groups such as Indigenous populations. However, despite the heavy burden of disease in Indigenous communities, there is scant data on immunity to influenza viruses in Indigenous populations from around the world.

As CD8^+^ T-cell recognition is determined by the spectrum of human leukocyte antigens (HLAs) expressed in any individual, and HLA profiles differ across ethnic groups, defining T-cell epitopes restricted by HLAs predominant in some Indigenous populations is necessary to understand pre-existing CD8^+^ T-cell immunity to influenza. We previously analyzed the HLA allele repertoire in Indigenous Australians^29^ and found that HLA-A*24:02 (referred to as HLA-A24 hereafter), an HLA associated with influenza-induced mortality during the 2009-pH1N1 outbreak^30^, is the second most prominent HLAs in Indigenous Australians^29,31^. HLA-A24 is also common to other Indigenous populations highly affected by influenza^17^. Thus, analysis of prominent influenza-specific CD8^+^ T-cell responses restricted by HLA-A24 is needed to understand the relationship between this allele and disease susceptibility. These specificities will also inform strategies to prime effective T-cell immunity in vulnerable communities. Here, we defined CD8^+^ T-cell immune landscapes against IAV and IBV, restricted by the mortality-associated HLA-A24 allomorph. We identified IAV- and IBV-specific HLA-A24 immunopeptidomes and screened immunogenicity of novel peptides in HLA-A24-expressing mice, peripheral blood of Indigenous and non-Indigenous HLA-A24^+^ healthy and influenza-infected individuals, and human tissues. Our studies provide evidence of the breadth of influenza-specific CD8^+^ T-cell specificities restricted by a mortality-associated risk allomorph HLA-A24. These findings have implications for the incorporation of key CD8^+^ T-cell targets in a T-cell-mediated vaccine to protect Indigenous people globally from unpredicted influenza viruses.

## RESULTS

### High prevalence of HLA-A24 expression in Indigenous populations

As HLA-A24 has been linked to pH1N1-related mortality^30^, we first determined HLA-A24 distribution in Indigenous and non-Indigenous populations worldwide using the published allele frequency database. Compared to the 10% global distribution of HLA-A24, the detected frequencies of HLA-A24 were the highest in Oceania (37%), North-East Asia (32.9%), Australia (21.4%) and Central and South America (20.6%) (**Fig. 1a**). This was mainly due to a particularly high HLA-A24 prevalence in Indigenous populations in those regions, especially in the Pacific. HLA-A24 was highly prevalent in Indigenous Taiwan Paiwan (96.1%), Papa New Guinea Karimui Plateau Pawaia (74.4%), New Caledonia (60.7%), Alaskan Yupik (58.1%), New Zealand Maori (38%), American Samoans (33%), Chile Easter Island (35.8%) and some Australian Indigenous people (24%) (**Fig. 1a**), which highlights its key importance in shaping CD8^+^ T-cell immunity in Indigenous populations.

**Figure 1.**
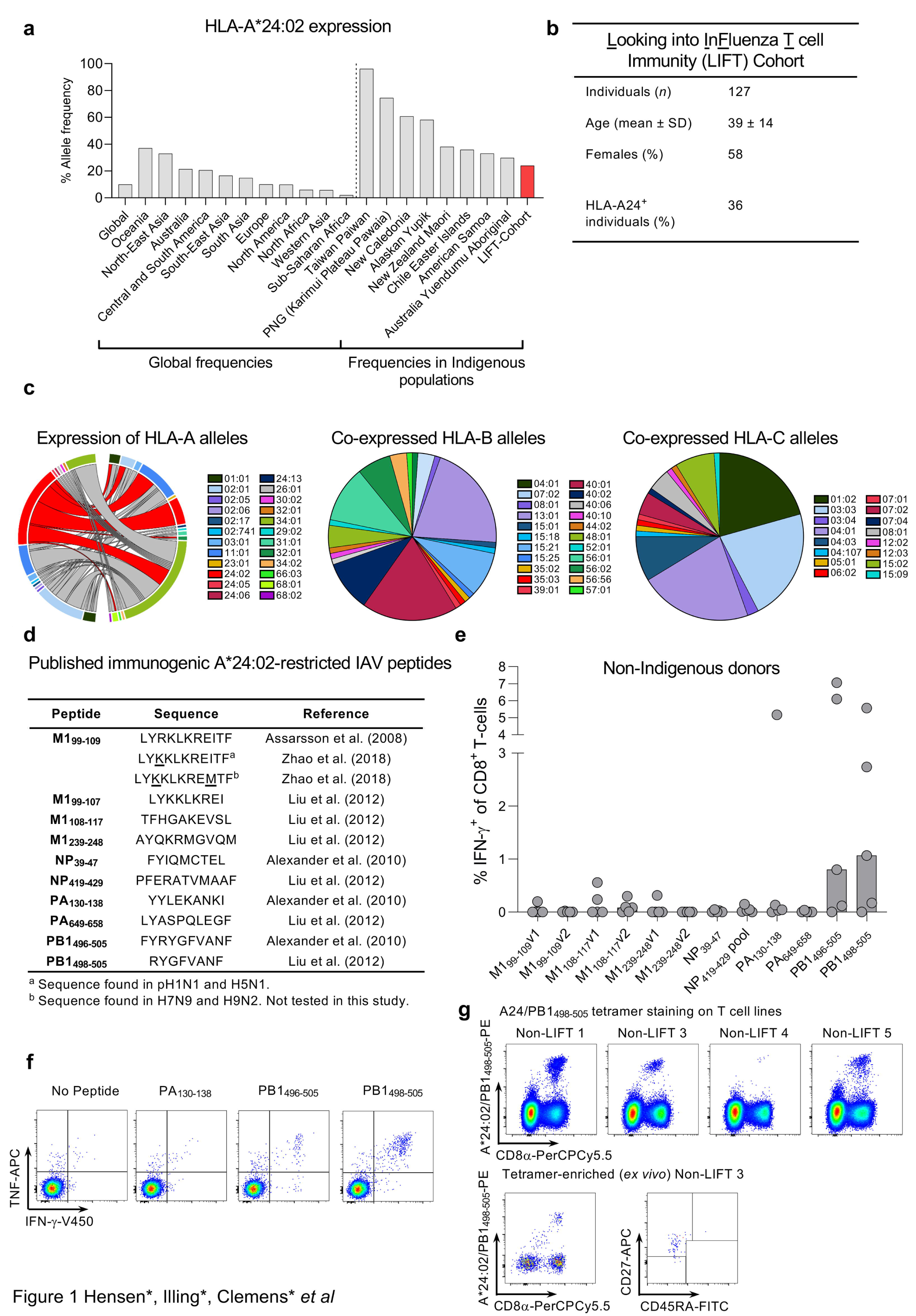
HLA-A24 expression and characterization of published IAV epitopes. **(a)** Allele frequency of HLA-A*24:02 according to geographic region (left panel) and Indigenous population in the pacific region (right panel) (source www.allelfrequencies.net accessed 20/01/2020). **(b)** Participant characteristics of the Looking into Influenza T-cell Immunity (LIFT) cohort. **(c)** Co-expression of HLA-A, B and C alleles with the HLA-A*24:02 allele in the LIFT cohort, where Circos plot is shown for HLA-A and pie charts are shown for HLA-C and B alleles that are co-expressed with the HLA-A*24:02 allele. (**d)** List of immunogenic HLA-A*24:02 IAV peptides from previous studies that were **(e)** screened for immunogenicity via IFN-γ-ICS assay following peptide-expansion of PBMCs from 5 non-Indigenous individuals (median bar graphs are shown). **(f)** Representative FACS plots of cytokine expression for immunogenic peptides. **(g)** A24/PB1_498-505_-tetramer staining of peptide-expanded T-cell lines (top panels) and after *ex vivo* TAME from non-LIFT 3 donor (bottom panels) showing the TAME-enriched population and phenotype.

To define influenza-specific CD8^+^ T-cell responses in Indigenous Australians, we recruited 127 participants from the Northern Territory, Australia into the LIFT (**<u>L</u>**ooking **<u>I</u>**nto in**f**luenza **<u>T</u>**-cell immunity) cohort^29^. The mean age of the participants was 39 years, with a standard deviation of 14 years and 58% of female participants. 36% of the LIFT donors expressed at least one HLA-A24 alleles, with 33% of those being HLA-A24 homozygous (**Fig. 1b)**. Notably, HLA-A24 was most commonly expressed with HLA-A*11:01, -A*34:01, -B*13:01, -B*15:21, -B*40:01, -B*40:02, -B*56:01, -C*04:03, -C*03:03, -C*04:01 and - C*04:03 in Indigenous Australians, and were less expressed with alleles common in Caucasian populations, such as HLA-A*01:01, -A*02:01, -B*07:02 and -B*08:01 (**Fig. 1c**).

### CD8^+^ T-cell responses towards published IAV-specific HLA-A24-epitopes

A handful of IAV-specific HLA-A24-restricted epitopes have been described^32–35^ (**Fig. 1d**). We aimed to validate the immunogenicity of these epitopes by probing memory CD8^+^ T-cells within peripheral blood mononuclear cells (PBMCs) of healthy non-Indigenous HLA-A24-expressing donors **(Supplementary Table 1)** using an *in vitro* peptide stimulation assay (**Fig. 1e-g)**. Only 3 out of 12 peptides (PB1_498-505_, PB1_496-505,_ PA_130-138_) induced CD8^+^ T-cell proliferation and IFN-γ/TNF production in a limited number of donors (3/5, 3/5 and 1/5, respectively) (**Fig. 1e,f**). We deduced that the minimal epitope for PB1_496-505_ responses came from the PB1_498-505_ epitope. The specificity of PB1_498-505_-specific CD8^+^ T-cell responses, observed in multiple donors, was further verified by A24/PB1_498-505_ tetramer staining on both *in vitro*-cultured A24/PB1_498-505_ CD8^+^ T-cell lines and A24/PB1 ^+^CD8^+^ T-cells detected directly *ex vivo* by tetramer enrichment (**Fig. 1g**). Thus, while 3 of the previously published peptides elicited IFN-γ responses in a selected number of HLA-A24-expressing individuals, we sought to determine whether as yet unidentified epitopes might provide more robust IAV-specific CD8^+^ T-cell responses in HLA-A24-expressing individuals.

### Identification of novel HLA-A24-restricted IAV and IBV epitopes

To identify new A24/influenza-derived epitopes, we utilized an immunopeptidomics approach to sequence HLA-bound peptides on influenza-infected cells by liquid chromatography with tandem mass spectrometry (LC-MS/MS)^27^. Experiments were performed with class I-reduced (CIR) B-lymphoblastoid cell line (minimal HLA-B*35:03 surface expression but normal levels of HLA-C*04:01 expression^36^) and an HLA-A*24:02-transfected CIR cell line (CIR.A24)^37^. Initial experiments were performed in both cell lines at 16 hours post-infection with and without the HKx31 IAV virus, isolating peptide-bound HLA-I molecules post-lysis utilizing the pan HLA-I antibody W6/32 (**Supplementary Data 1**). In subsequent experiments, to improve confidence of assignment of binding to HLA-A24, HLA-C*04:01, depletion of the lysates was performed with the HLA-C-specific antibody DT9 prior to isolation of the remaining HLA-I (A*24:02>>B*35:03) with W6/32 antibody. Utilizing this workflow^38^, we assessed peptide presentation in CIR.A24 cells at 2, 4, 8 and 12 hours post-infection with either A/HKx31 or B/Malaysia/2506/2004 viruses. Analyses were restricted to up to 12 hours post-infection due to observations of marked HLA-I downregulation at 16 hours post-infection^39^.

In total, 12 immunopeptidome data sets containing HLA-A24-restricted peptides were generated including 3 from uninfected CIR.A24 cells, 5 from HKx31-infected and 4 from B/Malaysia-infected CIR.A24 cells (**Supplementary Fig. 1a, Supplementary Data 1**). An additional 3 data sets for endogenous HLA-I of CIR cells (CIR W6/32 isolation of HLA-B*35:03 and HLA-C*04:01 after 16 hours HKx31 infection; CIR.A24 - DT9 isolation of HLA-C*04:01 from uninfected cells and after 12 hours HKx31 infection) and 2 data sets for endogenous HLA-II (CIR.A24: HLA-DR12, -DPB1*04:01,04:02 and -DQ7 from uninfected and 12 hours HKx31 infection) were also generated as comparators to help establish true HLA-A24 binders (**Supplementary Fig. 1b,c**). Comparisons to previously identified B/Malaysia-derived HLA ligands for CIR were also used^27^ to distinguish of HLA-B*35:03 and HLA-C*04:01 binding peptides from those binding to HLA-A*24:02. Across the 12 data sets, a total of 9051 non-redundant peptide sequences were assigned as HLA-I ligands using a 5% false discovery rate (FDR). As expected for HLA-I ligands, the majority of peptides were 9-11 amino acids in length but dominated by 9mers (**Fig. 2a**). Consistent with the HLA-A24 peptide-binding motif generated by NetMHC4.0^40,41^ motif viewer, enrichment of Tyr/Phe at P2 and Phe/Leu/Ile at P9 were observed (**Fig. 2b**). Peptides binding the endogenous HLA-I of CIR were not removed in this analysis due to the similar preference of HLA-C*04:01 for 9mer peptides possessing Phe/Tyr at P2 and Phe/Leu at P9 which may result in shared ligands (**Supplementary Fig. 1d**). To maximize identification of potential virus-derived peptides, assignments to the viral proteome or 6-frame translation of the viral genome were considered without a FDR cutoff. Instead, lack of appearance in uninfected data sets and predicted binding affinity (NetMHCpan4.0^42–44^) for HLA-A24 were used determine likely candidate epitopes. Thus, 52 HKx31-derived and 48 B/Malaysia-derived peptides were identified as potential HLA-A24-restricted epitopes (**Fig. 2c**), of which 26 IAV-derived and 29 IBV-derived peptides were identified at a 5% FDR **(Supplementary Data 1)**. The identified peptides spanned the viral proteomes including frame-shift proteins, representing 6 IAV proteins and 9 IBV proteins (**Fig. 2d,e**). Interestingly, most HKx31-derived ligands mapped to PB2>PB1>HA viral proteins and none were observed from NA or M1, while B/Malaysia-derived ligands predominantly mapped to NP/HA>NA. During the time course analyses, broadest peptide identification was achieved for both viruses between 8-12 hours post-infection, while no influenza-derived peptides were identified at 2 hours post-infection, and those identified at 4 hours were of lower confidence (**Fig. 2f,g, Supplementary Data 1**). 10 potential HKx31-derived ligands were also identified for each of HLA-B*35:03 and HLA-C*04:01 based on predicted binding and/or appearance in control data sets (**Supplementary Fig 1e, Supplementary data 1**). Furthermore, analysis of the peptides presented by HLA-II molecules showed domination of the virus-derived immunopeptidome by HA (**Supplementary Fig 1f, Supplementary Data 1**), as previously observed for IBV^27^. A selection of 48 IAV and 41 IBV peptides were synthesised for subsequent screening (Supplementary **Tables 2,3, Supplementary Data 1**). Notably, most synthetic peptides showed highly similar fragmentation patterns and retention times to the discovery data, supporting the original identifications (**Supplementary Data 1**).

**Figure 2.**
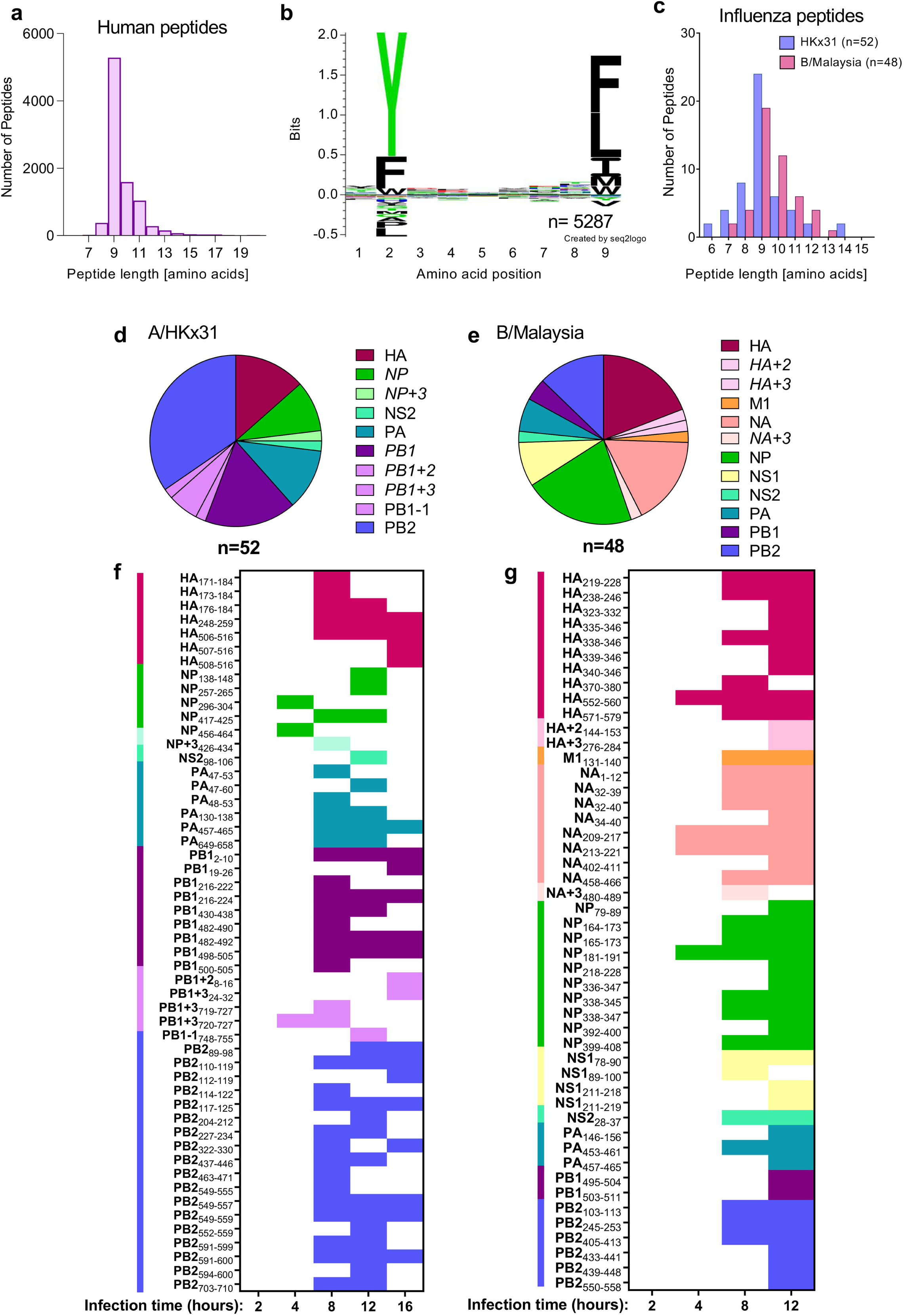
LC-MS/MS analyses of immunopeptidome from HLA-A*24:02^+^ influenza-infected cells reveal potential HLA-A*24:02-restricted T-cell epitopes. **(a)** Length distribution of human proteome-derived HLA-I ligands of CIR.A24 isolated using the pan HLA-I antibody w632. Numbers of non-redundant sequences of ≤ 20 amino acids identified at a 5 % FDR across the 12 experiments in which HLA class I was isolated from CIR.A24, filtered for peptides identified in HLA class II isolations at a 5% FDR. **(b)** Sequence logo derived from human 9mer peptides in **(a)** using Seq2logo2.0^69^. **(c)** Peptide length distribution of IAV (HKx31) and IBV (B/Malaysia) derived peptides identified as likely HLA-A*24:02 ligands in this study (no FDR cut-off applied) as shown in Supplementary Data 1. **(d,e)** Distribution of IAV-**(d)** and IBV-derived **(e)** A*24:02 ligands from **(c)** across the viral proteomes, including potential identifications from alternate reading frames (PB1+2, PB1+3, PB1-1, HA+2, HA+3 and NA+3). **(f,g)** Identification of specific ligands derived from IAV **(f)** and IBV **(g)** in isolations performed at 2, 4, 8, 12 and 16 hours (16 hours IAV only) post infection. Colored squares represent identification of each peptide derived from viral proteins as indicated in legend from **(d,e)**. In all panels, n=number of peptides.

### Broad CD8^+^ T-cell responses in IAV-infected HLA-A24 mice are biased towards PB1- and PB2-derived peptides

To determine the immunogenicity of novel IAV- and IBV-derived peptides during primary and secondary influenza virus infection *in vivo*, we utilized HLA-A24-expressing transgenic (HHD-A24) mice^45^. These mice are not confounded by prior exposure to infections nor co-expression of other competing major histocompatibility complex I (MHC-I) molecules, and thus provide a valuable tool for screening peptide antigens *in vivo*. For primary IAV analyses, 6-10 week old mice were intranasally (i.n.) infected with 200 pfu of H3N2 HKx31 virus, while secondary experiments were performed by an i.n. heterologous challenge with 200 pfu H1N1 PR8 6-8 weeks after the primary HKx31 infection. At day (d) 10 post-infection, spleen and bronchoalveolar lavage (BAL, combined from 4-5 mice) were stimulated with 54 peptides; including 48 novel peptides identified by mass-spectrometry and the previously published PB1496-505, M199-109, M1108-117, M1239-248, NP39-47 & NP419-429 peptides (**Supplementary Table 2**). The responses were detected by 5-hr *ex vivo* peptide stimulation and measurement of IFN-γ production by ICS. Our data revealed that A24/IAV-specific CD8^+^ T-cell responses were immunodominant (>5% IFN-γ^+^ of CD8^+^ T-cells) towards 2 PB1- and 3 PB2-derived peptides: PB1_216-224_ (mean of 10.2% IFNγ^+^ of CD8^+^ cells in spleen, 16.5% in BAL), the published PB1_498-505_ (14.8% spleen, 23.3% BAL), PB2_549-557_ (10.1% spleen, 24.9% BAL), PB2_549-559_ (11.1% spleen, 24.4% BAL) and PB2_463-471_ (6% spleen, 9.3% BAL) (**Fig. 3a**). Two additional subdominant (1%< IFN-γ^+^ of CD8^+^ T-cells <5%) CD8^+^ T-cell responses were directed towards PB2_322-320_ (3.9% spleen, 7.7% BAL) and previously published NP_39-47_ (2.1% spleen, 7.7% BAL), while marginal CD8^+^ T-cell responses were detected, mainly in BAL, towards 6 other peptides (HA_248-259_, PA_47-60_, PA_130-138_, PB1_2-10_, PB1500-505, PB2703-710).

**Figure 3.**
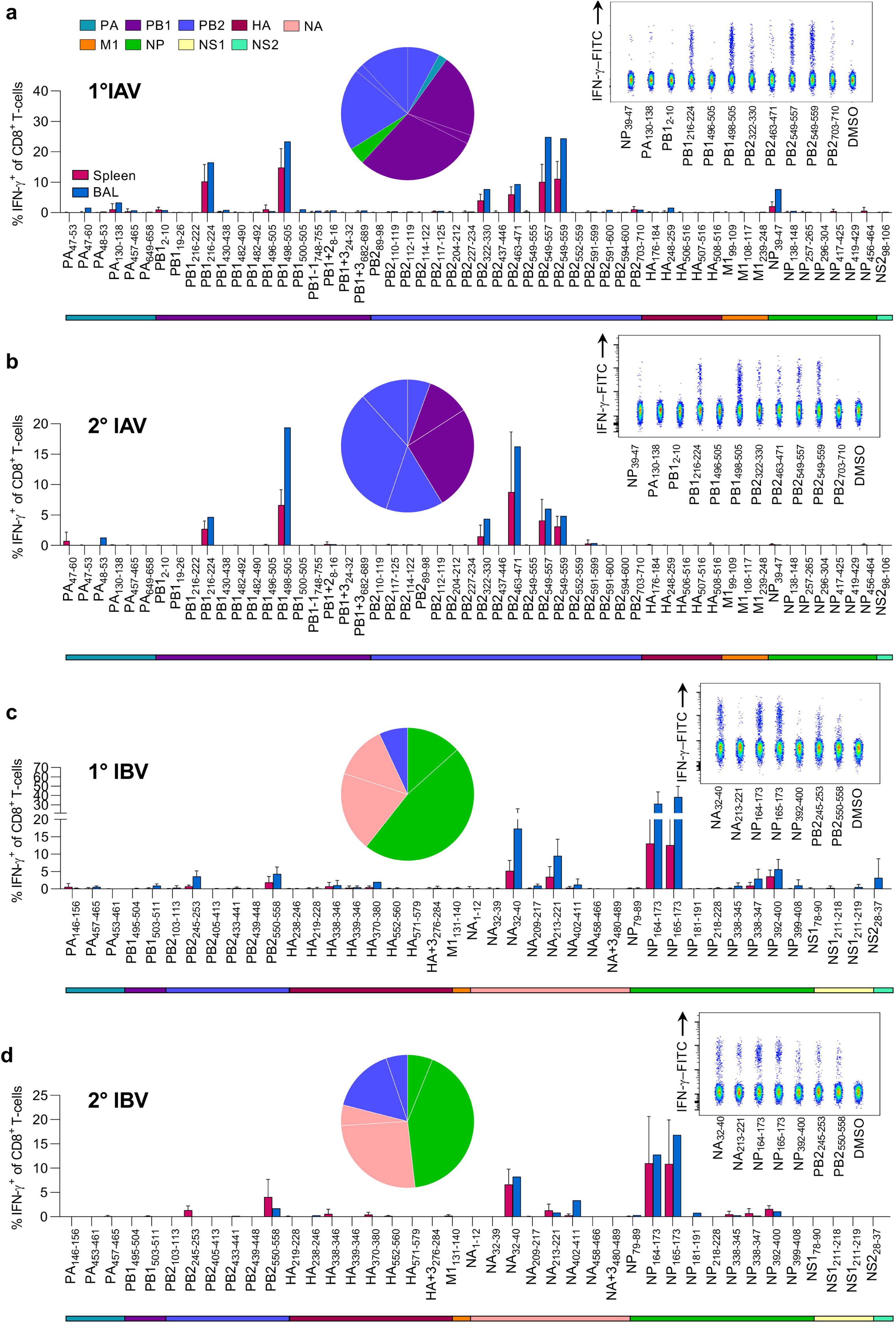
Screening of immunogenic IAV and IBV peptides in HHD-A24 mice. **(a,c)** Frequencies of IFN-γ^+^ CD8^+^ T-cells in spleen (individual mice) and BAL (pooled mice) after primary infection with either IAV (100 pfu/30μl i.n. A/HKx31) or IBV (200 pfu/30μl i.n. B/Malaysia/2506/04), respectively, from 6-10 week-old HLA-A24-expressing mice. Cells from spleen and BAL were isolated 10 days post-infection and restimulated with individual peptides (**a**, n=5-10 mice from 1-2 independent experiments) **(c**, n=9-14 mice from 2-3 experiments**). (b,d)** IFN-γ^+^ frequencies following a secondary heterologous challenge with IAV (200 pfu/30μl of A/PR8) and IBV (400 pfu/30μl of B/Phuket/3073/2013), 6 weeks after primary infection, respectively. Cells were isolated 8 days post-infection for ICS assay (b, n=5 mice from 1 experiment) (d, n=4 mice from 1 experiment). Bars indicate mean+SD for spleen and BAL for **(c)** only. Pie charts depict protein origin of immunogenic peptides and the average contribution to the total IFN-γ response in spleen for each primary and secondary IAV/IBV responses. Coloured bars below bar chart indicate protein origin of characterised peptides. Representative concatenated FACS plots from splenocytes of immunodominant and subdominant responses compared to DMSO background control are shown in the top-left panels.

During the acute phase (d8) of secondary challenge, CD8^+^ T-cell responses were similarly directed at 6 immunogenic peptides found in the primary infection (PB1_216-224_, PB1_498-505_, PB2_322-330_, PB2_463-471_, PB2_549-557_ & PB2_549-559_) (**Fig. 3b**). CD8^+^ T-cell responses towards the marginal epitopes observed in the primary infection were no longer detected. While the reason for the loss of HA_248-259_ could be explained by sequence variation between HKx31 and PR8 (IYWTIVKPGDVL vs. **<u>Y</u>**YWT**<u>L</u>**VKPGD**<u>T</u>**I) all internal proteins are shared between both viruses. Analysis of influenza-specific CD8^+^ T-cell numbers showed significant reductions in epitope-specific CD8^+^ T-cells for 9 out of 11 specificities (NP_39-47_, PA_130-138_, PB1_2-10_, PB1_216-224_, PB1_496-505_, PB1_498-505_, PB2_322-330_, PB2_703-710_ & PB2_549-559_) following secondary infection (**Supplementary Fig. 2**).

Thus, our *in vivo* screening in HHD-A24 mice identified 6 IAV derived immunogenic peptides during primary and secondary IAV infection, with prominent CD8^+^ T-cell responses being heavily biased towards PB1- and PB2-derived peptides **(Supplementary Table 2)**. This is of key importance as the current T-cell vaccines in clinical trials focus mainly on internal proteins like NP and M1^46–50^ which may be poorly immunogenic in HLA-A24-expressing individuals at risk of severe influenza disease.

### Prominent A24/CD8^+^ T-cell specificities in IBV infection

While the CD8^+^ T-cell responses towards IBV have been studied in detail for HLA-A*02:01-expressing individuals^27^, there remains a lack of known CD8^+^ T-cell epitopes for other HLAs. Here, we determined the immunogenicity of newly identified IBV-derived peptides using primary and secondary infection in HHD-A24 mice. HHD-A24 mice were i.n. infected with 200 pfu B/Malaysia/2506/2004. On d10, spleen and BAL were restimulated with individual peptides (**Supplementary Table 3**) to measure the corresponding CD8^+^ T-cell responses. Novel immunodominant IFN-γ-producing A24/CD8^+^ T-cell specificities were found against a range of IBV peptides including NP_164-173_ (mean of 13.1% in spleen, 31.15% in BAL), the shorter NP_165-173_ (12.6% spleen, 38.4% BAL) and NA_32-40_ (5.2% spleen, 17.3% BAL), while subdominant A24/CD8^+^ T-cell responses were detected towards an additional four IBV-derived peptides (NA_213-221_, NP_392-400_, PB2_245-253_, PB2_550-558_) (**Fig. 3c**). Interestingly, although A24/CD8^+^ T-cells could respond to the 10mer NP_164-173_ by peptide re-stimulation, peptide-loaded MHC-tetramers showed that these CD8^+^ T-cells bound only the 9mer NP_165-173_ (data not shown), suggesting that the minimal epitope was the 9mer.

To define IBV-specific CD8^+^ T-cell responses following the secondary challenge, B/Malaysia/2506/2004-infected mice were challenged i.n. after 6-8 weeks with 400 pfu B/Phuket/3073/2013. At d8 after challenge, IBV-specific CD8^+^ T-cell responses resembled those after primary IBV infection (**Fig. 3d**), but were focussed more towards the immunodominant epitopes, similar to IAV-specific A24/CD8^+^ T-cell responses following secondary challenge. Although the B/Phuket virus differs in one amino acid in the NP_164-173_/NP_165-173_ and NA_213-222_ restimulating with the B/Malaysia variants showed cross-reactivity of CD8^+^ T-cells between both variants. All other immunogenic epitopes were conserved between both strains. In contrast to IAV infection, the total number of epitope-specific CD8^+^ T-cells for all immunogenic epitopes after secondary challenge remained comparable in the spleen (**Supplementary Fig. 2**). Thus, our data identified prominent A24/CD8^+^ T-cell responses directed towards IBV during primary and secondary influenza virus infection in HHD-A24 mice (**Supplementary Table 3**).

### IAV-specific CD8^+^ T-cells in HLA-A24-expressing Indigenous and non-Indigenous individuals

Having identified prominent IAV-derived CD8^+^ T-cell epitopes towards the primary and secondary infection in HHD-A24 mice, it was of key importance to define immunodominant CD8^+^ T-cell sets in HLA-A24-expressing Indigenous and non-Indigenous individuals. For humans, different influenza strains of the same subtype are often co-circulating and mutating to create different variants of the same epitope region. In addition to our identified panel of HLA-A24-binding peptides, we searched the Influenza research database (https://www.ncbi.nlm.nih.gov/genomes/FLU/Database/nph-select.cgi?go=database) to include the most frequent virus strains circulating in South-East Asia and Australia and identified naturally occurring variants of our HLA-A24 binding peptides utilizing the “*Identify short peptides in proteins”* analysis from Influenza research database (fludb.org) (n=61-3877 sequences dependent on protein) to include in epitope mapping (**Supplementary Table 4**). We probed memory CD8^+^ T-cell populations, by firstly stimulating PBMCs from healthy Indigenous HLA-A24-expressing donors with 5 IAV peptide pools for 13 days. Each pool contained 7-13 peptides (**Supplementary Table 2**), of which each variant (**Supplementary Table 4**) was always included in the same pool as the wildtype peptide identified in the immunopeptidome studies. We observed CD8^+^ T-cell responses towards pools 1 and 2 via an IFN-γ/TNF ICS assay (**Fig. 4a**), and subsequently cell cultures from those pools were restimulated with individual peptides (+variants) to map the immunogenic epitopes. In 5/5 Indigenous donors tested, CD8^+^ T-cell responses were dominated towards PB1_498-505_ (median of 0.93% IFN-γ of CD8^+^ T-cells; also immunodominant in mice **Fig. 3ab**), its longer 10mer version PB1_496-505_ (median of 0.64%; detected marginally after 1° IAV infection in mice), PA_649-658_ (median of 0.73%; not detected in mice) and NP_39-47_ (median 0.16%, immunogenic following 1° IAV infection in mice). Another 6 subdominant HLA-A24-restricted epitopes were identified in a select number of donors (PB2_110-119_ in 4/5 donors, NS2_98-106_ in 3/5, PB1_216-224_ in 3/5, PB2_549-557_ in 1/5, HA_176-184_ in 3/5 and PB2_703-710_ in 3/5), of which PB1_216-224_, PB2_549-557_ and PB2_703-710_ were also detected in HHD-A24 mice. Interestingly, only 2/4 immunodominant epitopes observed in the Indigenous donors elicited comparably robust responses in 5 non-Indigenous donors screened (published epitopes: PB1_498-505_ median 0.93% vs 1.1%, PB1_496-505_ median 0.64% vs 0.8%), while the other two epitopes, PA_649-658_ and NP_39-47_ were poorly immunogenic in non-Indigenous donors who instead responded well to the PB2_549-557_ epitope, absent in 4/5 Indigenous donors. Such differential epitope preference and immunodominance hierarchies between Indigenous and non-Indigenous donors is perhaps influenced by different HLA co-expressions or infection history.

**Figure 4.**
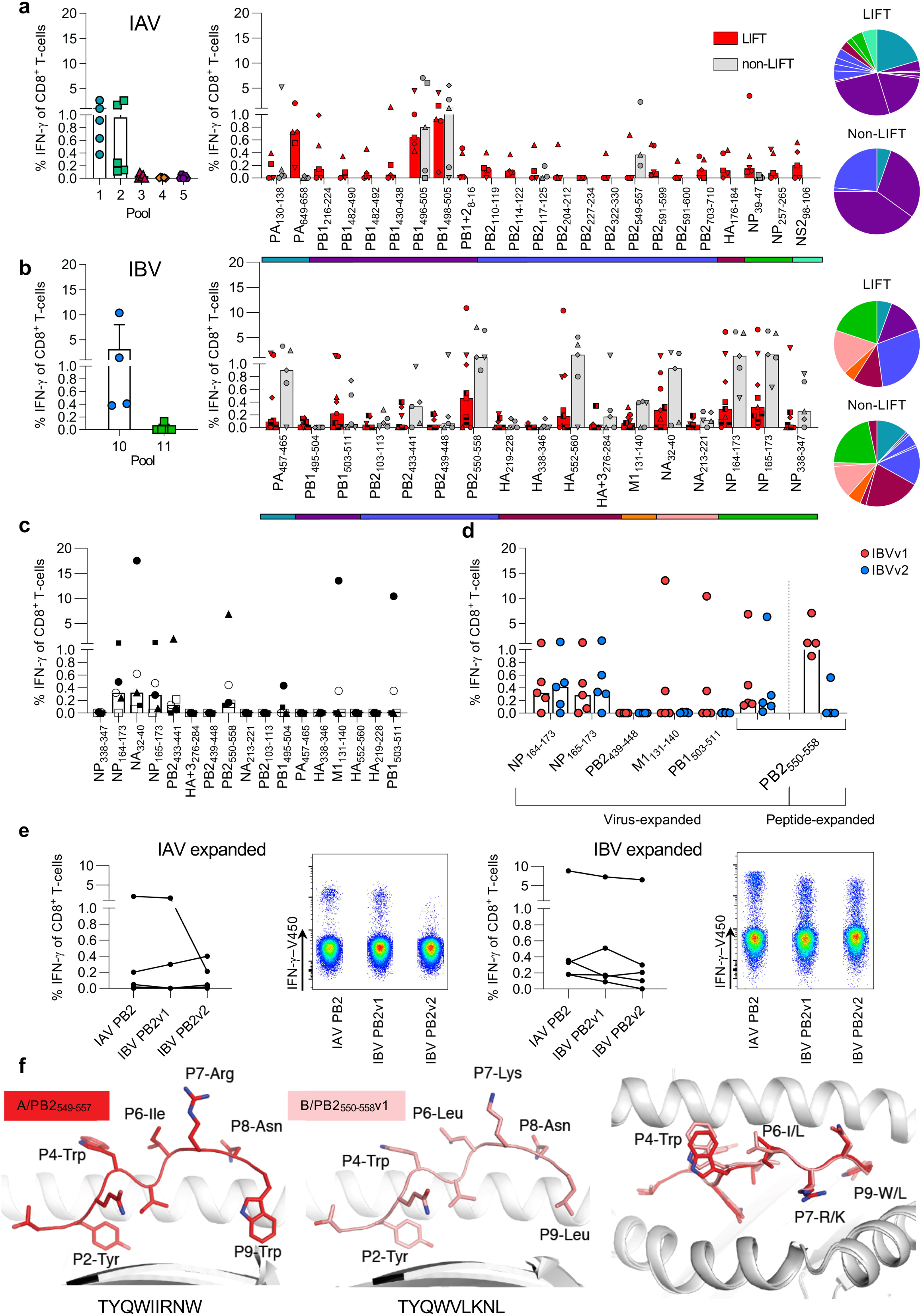
*In-vitro* screening for immunogenicity of IAV and IBV epitopes in human PBMC. **(a,b)** Frequencies of IFN-γ^+^CD8^+^ T-cells after PBMCs were expanded with IAV **(a)** or IBV **(b)** peptide pools for 13 days and restimulated with CIR.A24 cells pulsed with the corresponding peptide pools (left panel). Right panels show dissection of each peptide from IAV pools 1 and 2 **(a)** or IBV pool 10 **(b)** using single peptide-pulsed CIR.A24 cells after 15 days of peptide-pool T-cell expansions in LIFT and non-LIFT. **(c,d)** Individual IBV peptide IFN-γ^+^CD8^+^ T-cell responses in non-LIFT donors following expansion with B/Malaysia/2506/04-infected CIR.A24 cells for 15 days. For the ICS, peptides derived from B/Malaysia/2506/04 **(c)** or a comparison between autologous (v1) and the alternative circulating variant (v2) was used. Symbols indicate individual donors screened. **(d)** IBV PB2_550-558_ v1 and v2 responses after virus expansion versus IBV PB2_550-558_ v1 peptide expansion are shown on the right (d). **(a-d)** Bars indicate median. **(e)** Cross-reactive PB2 responses of IAV PB2, IBV PB2 v1 and IBV PB2v2 peptides following PBMC expansion with IAV (A/HKx31) (left panels) or IBV (B/Malaysia/2506/04) infected (right panels) CIR.A24 cells from 5 non-LIFT donors with representative concatenated FACS plots. **(c,d)** Pie charts depict protein origin of immunogenic peptides and the median contribution to the total IFN-γ response for Indigenous (LIFT) and non-Indigenous (non-LIFT) donors. Coloured bars below bar chart indicate protein origin of characterised peptides. **(f)** Crystal structures of HLA-A*24:02 presenting the PB2 peptide variants IAV PB2 (red, left) and IBV PB2v1 (pink, middle), and an overlay of both peptides (right).

### Breadth of A24/CD8^+^ T-cell specificities towards IBV in Indigenous and non-Indigenous donors

Following identification of novel IBV-derived peptides in HHD-A24 mice, we defined CD8^+^ T-cell responses towards IBV peptide pools, comprising 41 peptides identified by mass spectrometry (**Supplementary Table 3**) and an additional 14 variants (**Supplementary Table 4**), in HLA-A24-expressing Indigenous and non-Indigenous donors. In accordance with the HHD-A24 mouse data, we found broad A24/CD8^+^ T-cell responses directed towards pool 10, which mapped 6 major immunogenic epitopes spanning 5 different proteins (NP_164-173_/NP_165-173_, NA_32-40_, PB2_550-558_, PA_457-465_, HA_552-560_ and PB1_503-511_) (**Fig. 4b**). CD8+ T-cell responses to these peptides were found in 7/9, 8/9, 6/9, 6/9, 5/9 and 5/9 of Indigenous and 5/5, 5/5, 4/5, 5/5 and 4/5 of non-Indigenous donors, respectively, with comparable IFNγ^+^CD8^+^ T-cell frequencies between Indigenous and non-Indigenous donors. As our experiments examined the immunogenicity of A24/CD8^+^ T-cells following *in vitro* peptide-driven expansion, we further verified these novel IBV epitopes in non-Indigenous donors by incubating HLA-A24^+^ PBMCs with IBV-infected (B/Malaysia/2505/2004) CIR.A24 target-cells for 15 days before dissecting individual peptide (+variant) responses from IBV pool 10. This confirmed the immunogenicity of at least 3 epitopes previously detected by peptide-driven expansions (NP_164-173_/NP_165-173_, NA_32-40_ & PB2_550-558_) (**Fig. 4c**). Interestingly, the same epitopes were observed in HHD-A24 mice after IBV infection (**Fig. 3c.d**).

Collectively, out of 41 newly identified HLA-A24-binding IBV epitopes by immunopeptidomics, we confirmed a total of 9 (22%) immunogenic epitopes after screening HLA-A24-expressing mice and humans. Of these, 3 were found in both humans and mice (NP_164-173_/NP_165-173_, NA_32-40_, PB2_550-558_), 3 were only found in humans (HA_552-560_, PA_457-465_ and PB1_503-511_), and the remaining 3 were only found in mice (PB2_245-253_, NA_213-221_ & NP_392-400_).

### A24/CD8^+^ T-cell cross-reactivity between IAV and IBV towards PB2 epitopes

The B/Malaysia/2505/2004 virus used here is from the Victoria (Vic) lineage, however, there is another IBV lineage that commonly co-circulates and infects humans called the Yamagata (Yam) lineage. To determine the potential for HLA-A24/CD8^+^ T-cell cross-reactivity across both IBV lineages, B/Malaysia/2505/2004 (Vic) virus-expanded CD8^+^ T-cells were restimulated with IBV peptide variants from the alternate Yam lineage. Cross-reactivity was demonstrated towards the NP_165-173_ epitope (YFSPIRVTF variant 1 (Vic only)), in which B/Malaysia/2505/2004-expanded CD8^+^ T-cells responded towards the Yam variant (YFSPIR**<u>I</u>**TF v2 (Yam only)) following ICS assay (**Fig. 4d**). Likewise cross-reactivity between Vic lineage and a variant found in the Yam lineage was also observed with the PB2_550-558_ variants (TYQWVLKNL (variant1, both Vic and Yam); TYQWVMKNL (variant 2, Yam only)) in the virus-expansion system, but 3/4 donors did only respond to v1 after peptide expansion (**Fig. 4d**).

We have previously reported cross-reactivity towards IAV and IBV (as well as ICV) in the HLA-A2 model with a single epitope sequence^27^. Here, none of the identified HLA-A24 IAV and IBV epitopes showed 100% sequence identity between strains. Instead, we identified a potential HLA-A24-restricted IAV/IBV cross-reactive candidate, the immunogenic IAV PB2_549-557_ TYQW**<u>IIR</u>**N**<u>W</u>** epitope. This epitope shares 55% amino acid identity with the cross-reactive IBV PB2_550-558_ variants. Indeed, virus-expansion with IAV/HKx31-infected CIR.A24 cells induced cognate IAV/PB2_549-557_^+^ (A/PB2_549-557_)CD8^+^ T-cell responses in 2/4 donors as well as cross-reactive responses to the IBV PB2_550-558_v1 (B/PB2_550-558v1_) variant and the B/PB2_550-558_v2 variant (**Fig. 4e**) The superiority of IBV at inducing cross-reactive CD8^+^ T-cells in contrast to IAV was validated with the reverse experiment, where virus-expansion with B/Malaysia/2506/2004-infected CIR.A24 cells induced highly robust CD8^+^ T-cell responses in 4/4 donors, capable of cross-recognizing all three different epitopes (**Fig. 4e**).

As we observed T-cell cross-reactivity between the A/PB2_549-557_ and B/PB2_550-558_ (**Fig. 4e**) as well as T-cell reactivity in transgenic mice for the overlapping peptide A/PB2_549-559,_ we next determined the impact of those variations within the PB2 derived peptides. We solved the structures of A/PB2_549-557_, the overlapping A/PB2_549-559_ and B/PB2_550-558_ peptides in complex with the HLA-A24 molecules at a resolution of 2.90 Å, 2.95 Å and 2.16 Å, respectively (**Supplementary Table 5**) with clear electron density for each peptide (**Supplementary Fig. 5**).

The 9mer A/PB2_549-557_ peptide adopted a canonical extended conformation within the cleft of HLA-A24, with anchor residues at P2-Tyr and P9-Trp, and a secondary anchor residue at P5-Ile. Solvent exposed residues were at P4-Trp, P6-Ile, P7-Arg and P8-Asn, representing a large surface accessible for TCR interaction. The P9-Trp of the peptide formed a network of interactions with HLA-A24 tyrosine residues at positions 116, 118 and 123 as well as the Leu95 (**Supplementary Fig. 3a-c**), likely assisting with stabilizing the complex reflected in the higher stability observed for the HLA-A24-A/PB2_549-557_ complex than with other peptides (**Supplementary Table 5**).

The B/PB2_550-558_ peptide differed to the A/PB2_549-557_ peptide at positions 5 (Ile to Val), 6 (Ile to Leu), 7 (Arg to Lys) and 9 (Trp to Leu) (**Fig. 4f**). Both peptides shared the same anchor residue at P2-Tyr and similar solvent exposed residues (except for P7-Lys) but differed at PΩ (P9). As Leu possessed a shorter side chain than Tyr at PΩ, the IBV peptide was not buried as deeply into the F pocket, which may explain the lower stability observed for the B/PB2_550-558_ peptide (Tm of 57°C) compared to A/PB2_549-557_ (Tm of 62°C) (**Supplementary Table 5**). Structural overlay of HLA-A24 presenting the A/PB2_549-557_ and B/PB2_550-558_ peptides showed that the antigen-binding cleft and both peptides adopted a similar conformation with an average root mean square deviation (r.m.s.d.) of 0.31 Å and 0.37 Å, respectively (for Cα atoms) (**Fig. 4f**), consistent with T-cell cross-reactivity observed towards these two peptides.

Although the 11mer A/PB2_549-559_ generated similar responses to the 9mer A/PB2_549-557_ in HHD-A24 mice (**Fig. 3a,b**), it was not immunogenic in peptide-pool screening in humans (**Supplementary Table 2** Pool 4)**(Fig. 4a)** as perhaps the minimal 9mer epitope was not exposed for T-cell recognition, due to the two additional C-terminal residues (P10-Glu and P11-Thr) (**Supplementary Fig. 3a,b**). Similar to the 9mer peptide conformation P2-Tyr and P9-Trp of the 11mer PB2_549-559_ act as primary anchor residues buried in the HLA-A24 antigen-binding cleft with the structural overlay of the peptides showing an r.m.s.d. of 0.48 Å (**Supplementary Fig. 3a,b**).Strikingly, the extra P10-Glu and P11-Thr residues of the 11mer extended outside the antigen-binding cleft, creating an unusual conformation that disturbed the interaction between the peptide and the HLA-A24 Lys146 at the C-terminal of the cleft. The Lys146 residue is a conserved residue in HLA molecules that helps stabilise the pHLA complex^51^. In the 9mer PB2_549-557_ peptide, Lys146 interacts with the carboxylic group of the PΩ residue (**Supplementary Fig. 3d**), however this interaction is lost in the 11mer due to the presence of the extra two residues P10-Glu and P11-Thr. (**Supplementary Fig. 3e**), thereby likely decreasing the pHLA stability compared to the shorter A/PB2_549-557_ peptide (**Supplementary Table 5**). Thus, the bulged conformation of the extra residues in the A/PB2_549-559_ may represent a challenge for TCRs interacting with the C-terminal end of the peptide. In compliance with the *in vitro* data, the structural data support the potential cross-reactivity of CD8^+^ T-cells between the A/PB2_549-557_ and the B/PB2_550-558_, verifying our previous findings of broad CD8^+^ T-cell immunity against influenza virus infections.

### HLA-A24 presents overlapping NP_164-173_ and NP_165-173_ peptides in different conformations

We observed robust comparable mouse (**Fig. 3c,d**) and human (**Fig. 4b**) responses to the overlapping IBV NP_164-173_ and NP_165-173_ peptides. This suggested that the 9mer NP_165-173_ peptide would be the minimal epitope and both peptides might present a similar core confirmation recognized by T-cells. To determine this possibility, we solved the structures of the overlapping B/NP_164-173_ and B/NP_165-173_ peptides in complex with HLA-A24 at a resolution of 2.75 and 1.51Å, respectively (**Supplementary Fig. 4,5g-j, Supplementary Table 5**). The B/NP_164-173_ and B/NP_165-173_ peptides adopted a canonical extended conformation in the cleft of HLA-A24 molecule (**Supplementary Fig. 4**). P2-Phe and P9-Phe anchor residues of the 9mer B/NP_165-173_ were buried deep inside the hydrophobic B and F pockets of the HLA (**Supplementary Fig. 4a**). Three residues were exposed to the solvent for possible TCR recognition (P1-Tyr, P6-Arg, P8-Thr). The P5-Ile and P7-Val of this 9mer peptide were partially-buried (**Supplementary Fig. 4e**).

Structural overlay of 9mer and 10mer B/NP peptides were different due to the 10mer’s extra residue at the N-terminus (r.m.s.d. of 1.36 Å), which shifted the anchor residues (**Supplementary Fig. 4b,c**). The substitution of P2-Tyr (NP_164-173_) for P2-Phe (NP_165-173_) occurred without major structural rearrangement, as both residues were large aromatic residues filling the B pocket (**Supplementary Fig. 4f**). However, the additional residue changed the secondary anchor residue at P3 from a small P3-Ser (NP_165-173_) to a large P3-Phe (NP_164-173_) (**Supplementary Fig. 4g**). The larger P3-Phe might stabilize the B pocket of the HLA-A24 better than the small P3-Ser, and therefore could explain the 7°C higher Tm observed for the NP_164-173_ than the NP_165-173_ in complex with HLA-A24 (**Supplementary Table 5**), which could also reflect the immunogenicity of this peptide. The largest structural difference between the two peptides was observed at the centre of the peptide (P6/7-Arg) with a maximum displacement of 3.9 Å for the Cα atom (**Supplementary Fig. 4d**). The P7-Arg of the NP_164-173_ peptide sat higher than the backbone of the NP_165-173_ peptide (**Supplementary Fig. 4c**) and was a prominent feature for potential TCR interaction of the 10mer peptide, contrasting with the hydrophobic nature of the 9mer NP_165-173_ peptide. Thus, the structures of HLA-A24 presenting the two NP peptides showed that, despite being overlapping peptides that differ only by one residue, the NP_164-173_ and NP_165-173_ peptides adopt different structural conformations. As a result, both peptides exposed different residues to the solvent, and hence would most likely be recognized by different TCRαβ repertoires.

### Protective capacity of novel HLA-A24-restricted IBV-derived CD8^+^ T cell peptides during *in vivo* infection of HHD-A24 mice

To determine the protective capacity of the novel CD8^+^ T cell epitopes in HHD-A24 mice, we performed a proof of principle experiment and vaccinated mice with 3 immunogenic IBV peptides (NP_164_, NP_392_, NA_32_) using a well-established prime/boost approach^27^, then infected mice i.n. with 1×10^3^ pfu B/Malaysia (**Fig. 5a**). Vaccination with HLA-A24-restricted peptides resulted in significant protection against IBV. This was shown by decreased disease severity on d4, d5 and d6 after IBV infection as measured by the body weight loss (**Fig. 5b**; p<0.05) as well as a significant ∼89% reduction in viral titers in the lung on d7 after IBV infection when compared to the mock-immunized group (p<0.05) (**Fig. 5c**). Additionally, there was a significant decrease (p<0.05) in the levels of inflammatory cytokines (MIP-1β, MIP-1a, RANTES) in d7 BAL of peptide-vaccinated mice in comparison to the mock-immunised animals (**Fig. 5d**). Thus, CD8^+^ T cells directed at our novel HLA-A24-restricted IBV-specific epitopes provide a substantial level of protection against influenza disease, as they can markedly decrease body weight loss, accelerate viral clearance and reduce the cytokine storm at the site of infection.

**Figure 5.**
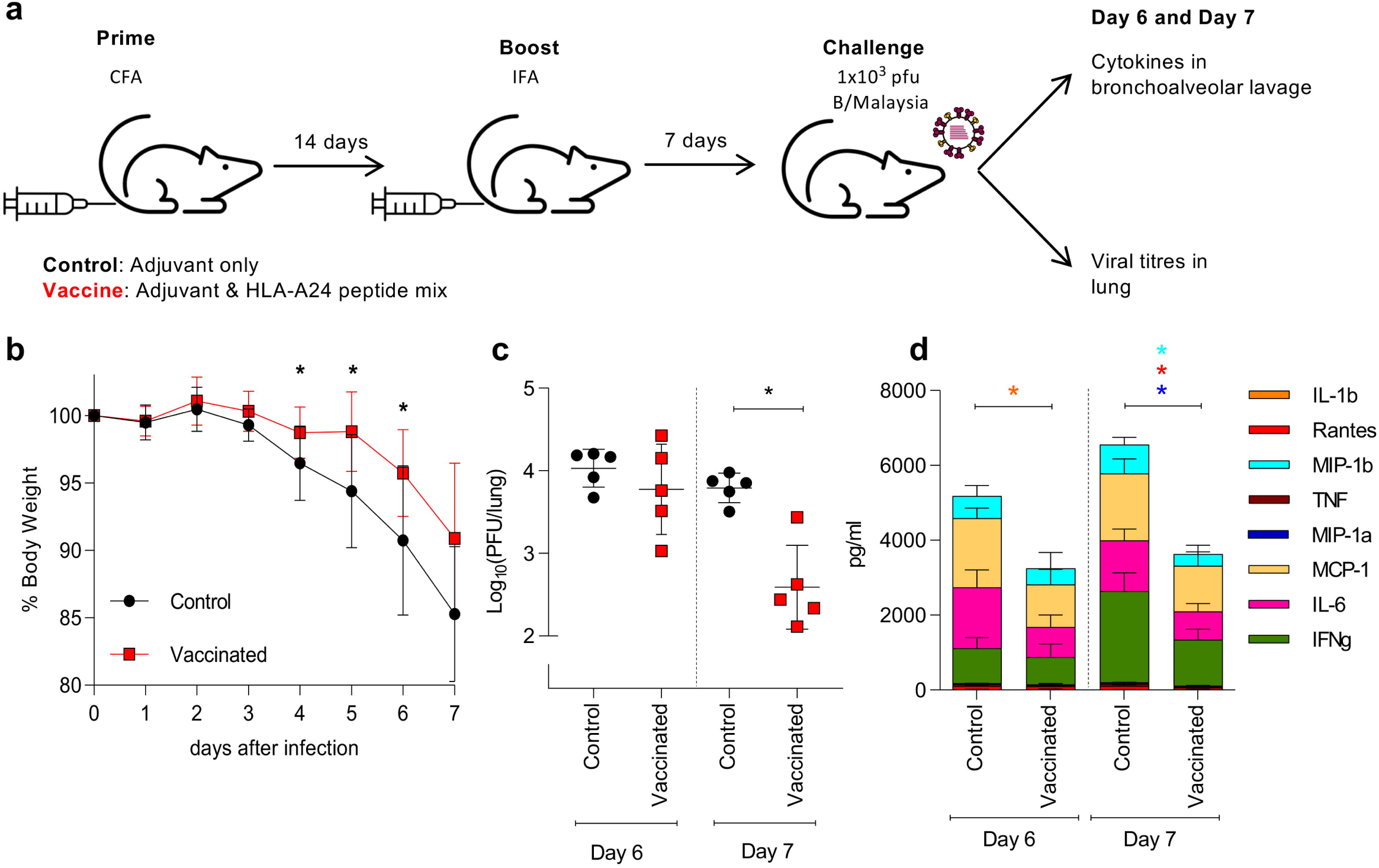
Protection of HDD.24 mice by vaccination with novel immunogenic A24 peptides. **(a)** Mice were vaccinated with 3 immunogenic IBV peptides (NP_164_, NP_392_, NA_32_) in adjuvant CFA or CFA alone (control). 14 days post-priming, immune responses were boosted with peptides/IFA or IFA alone. 7 days after the boost, mice were challenged with high dose of 1×10^3^ pfu B/Malaysia. Bronchoalveolar lavage and lungs were collected on day 6 and 7 post infection. **(b)** % body weight of mice after virus challenge at different days after infection. **(c)** Virus titre from homogenized lungs on day 6 and 7 post challenge. **(d)** Cytokine levels in BAL on day 6 and 7. Coloured stars indicate significant differences between control and peptide vaccinated groups. **(b-d)** Statistical significance was calculated using unpaired two-tailed t-test with *p<0.05.

### A24/CD8^+^ T-cell responses in IAV- and IBV-infected patient blood and healthy human tissues

Having identified the prominent IAV and IBV CD8^+^ T-cell specificities for Indigenous and non-Indigenous HLA-A24^+^-individuals, we sought to determine whether CD8^+^ T-cells specific for our newly identified epitopes were recruited and activated during acute influenza virus infection. We generated peptide/HLA-A24-tetramers to the most immunogenic IAV (A/PB1_498-505_) and newly characterised IBV epitopes (NP_165-173_ and NA_32-40_). These reagents allow direct *ex vivo* detection of IAV- and IBV-specific CD8^+^ T-cells using tetramer-associated magnetic enrichment (TAME)^52^ in both healthy and influenza-infected individuals (**Fig. 6a**, left panels). In healthy non-Indigenous and Indigenous donors, *ex vivo* mean precursor frequencies for A/PB1_498-505_^+^ and B/NP ^+^ CD8^+^ T-cells, were 4×10^−5^ and 1×10^−5^ of CD8^+^ T-cells, respectively (**Fig. 6b**). Non-Indigenous B/NA_32_^+^ frequencies were 1.8-9×10^−5^ of CD8^+^ T-cells. All tetramer^+^ frequencies fell within the range of previously published frequencies for memory IAV- or EBV-specific CD8^+^ T-cells^52,53^.

**Figure 6.**
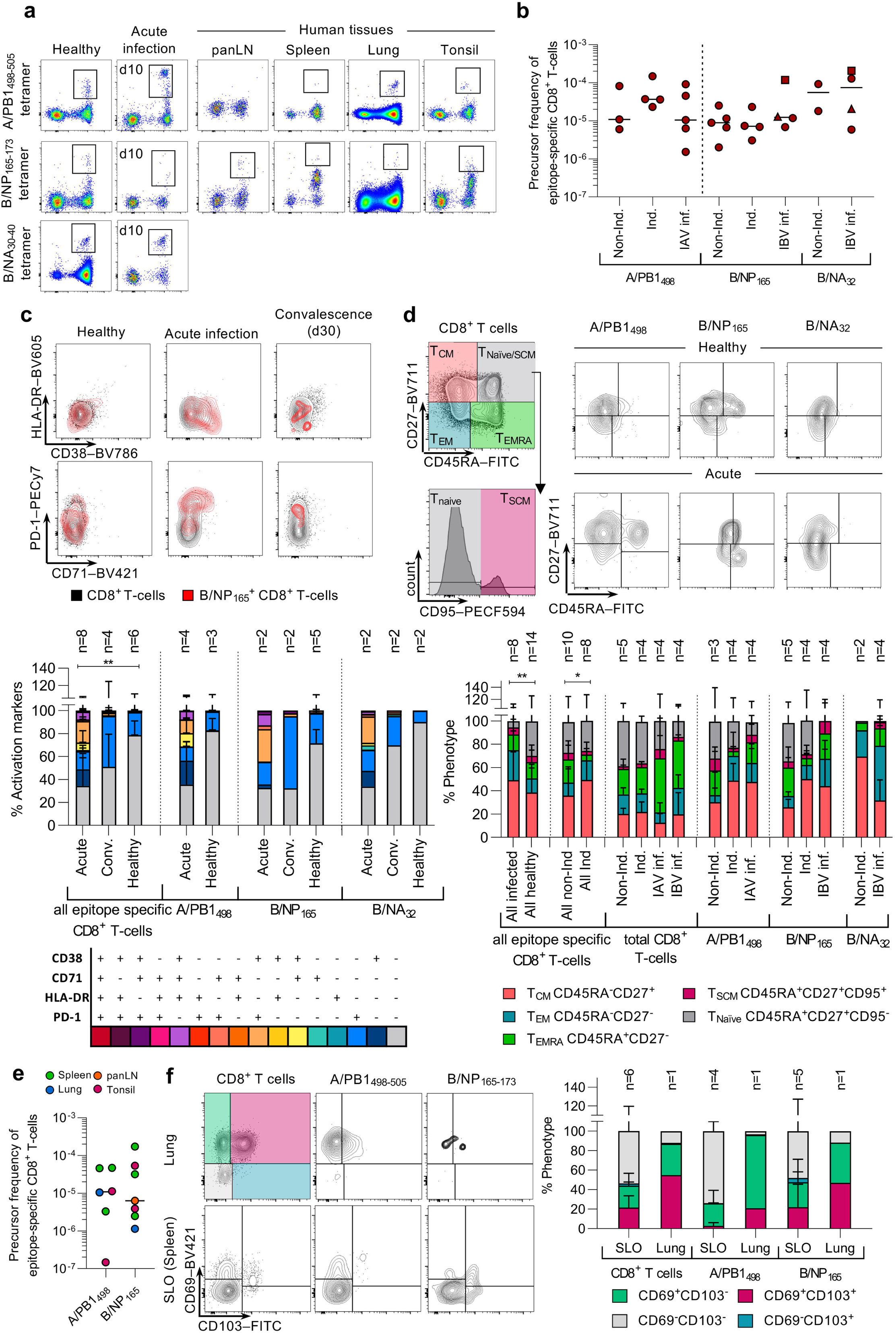
*Ex vivo* IAV- and IBV-specific CD8^+^ T-cells detected in HLA-A24^+^ PBMCs and tissues. **(a)** Tetramer-positive cells of three different specificities (A/PB1_498-505_, B/NP_165-173_, B/NA_32-40_) were enriched in human blood and tissues. Representative FACS plots of enriched fractions are shown for healthy donors, acute influenza-infected patients and tissues from deceased organ donors. **(b)** Tetramer^+^ precursor frequencies in the blood of healthy LIFT and non-LIFT, and influenza-infected donors where acute and convalescent time-points are shown together (IBV-inf). Samples from donor IBV2-inf were collected at acute square and convalescent timepoint triangle. **(c)** Activation status of epitope-specific CD8^+^ T-cells in healthy, acute and convalescent donors. **(d)** T-cell differentiation phenotype of epitope-specific cells in healthy LIFT and non-LIFT donors, and influenza-infected patients. **(e)** Tetramer^+^ precursor frequencies of epitope-specific cells in human tissues. **(f)** CD103 and CD69 expression on epitope-specific cells in the lung compared to secondary lymphoid organs (SLO). **(b,e)** Lines represent the median. **(c,d, f)** Bars represent mean+SD.

Interestingly, as per our analysis in HLA-A*02:01-positive influenza patients^27^, the frequencies of A/PB1_498_^+^, B/NP_165_^+^ and B/NA_32_^+^ CD8^+^ T-cells in blood of IAV- and IBV-infected patients during infection were comparable to that of memory CD8^+^ T-cell frequencies (**Fig. 6b**) This, most likely reflects accumulation of influenza-specific CD8^+^ T-cells at the site of infection rather than in the peripheral blood. However, despite similar frequencies, their activation profiles varied greatly, as assessed directly *ex vivo* by expression of the activation markers CD71, HLA-DR, CD38, PD-1, and phenotypic markers CD27 and CD45RA (**Figure 6c,d**). In patients infected with IAV and IBV, tetramer^+^CD8^+^ T-cells displayed significantly (*p*<0.05) higher frequencies cells expressing activation markers, especially CD38^+^PD-1^+^ (18.7% vs. 0.8%), CD71^+^PD-1^+^ and CD38^+^ (14.6% vs. 0.1%), CD38^+^CD71^+^PD-1^+^ (6.7% vs 0.0%) and CD38^+^CD71^+^ (6.3% vs. 0.0%),, in comparison to epitope specific CD8^+^ T-cells of healthy donors (**Figure 6c**). In convalescent donors the dominant activation factor expressed in the epitope-specific CD8^+^ T cells was PD-1 (44.2% in convalescent vs. 18.4% in healthy). Phenotypic analysis of CD45RA, CD27 and CD95 expression confirmed these results and showed higher frequencies of CD45RA^-^CD27^-^ effector memory-like T-cells in IAV- and IBV-infected patients (mean of 25.9% vs. 12.2% in healthy, p=0.04), higher but difference in CD45RA^-^CD27^+^ central memory-like T-cells (49.1% vs. 38.5% in healthy, p=0.24), and less naïve T-cells (5.4% vs. 29.7% in healthy, p<0.05) compared to healthy individuals within the pooled IAV/IBV tetramer specificities (**Figure 6d**). Comparing all healthy donors, we observed a lower frequency of CD45RA^+^CD27^-^ T_EMRA_-like T-cells in epitope specific CD8^+^ T-cells from Indigenous compared to non-Indigenous donors (5.0% vs 20.0% in non-Indigenous, p<0.05) (**Figure 6d**). This difference was not observed in the total non-specific CD8^+^ T-cell population.

Importantly, HLA-A24-restricted influenza-specific CD8^+^ T-cells against A/PB1_498_ and A/NP_165_ were detected in multiple healthy human tissues directly *ex vivo* (**Fig. 6a**, right panels). A/PB1_498_-specific CD8^+^ T-cells were detected in the lung, spleen and tonsil at frequencies ranging 1.5×10^−7^ to 4.5×10^−5^ of total CD8^+^ T-cells (n=6 data points), but was not detected in the pancreatic lymph node (panLN) of one donor that had detectable A/PB1_498-505_-specific CD8^+^ T-cells in the spleen (**Fig. 6e; data not shown**). B/NP_165_-specific CD8^+^ T-cells were found across all the tissues (lung, spleen, tonsil, panLN, n=7 data points) at frequencies between 1.1×10^−6^ to 1.7×10^−4^. In human lung, IAV/IBV-specific CD8^+^ T-cells had large populations of CD69^-^CD103^+^ and CD69^+^CD103^+^ tissue-resident memory (T_RM_) T-cells (A/PB1_498-505_: 75% and 21%, B/NP_165-173_: 41% and 47% of tetramer-specific CD8^+^ T-cells, respectively) (**Fig. 6f**). Secondary lymphoid organs (SLOs) were predominantly CD69^-^ CD103^-^ circulating effector memory cells (range 23.8-85.7%).

Our findings demonstrate the presence of highly activated influenza-specific CD8^+^ T-cells against the published A/PB1_498_ epitope and the IBV epitopes identified here in HLA-A24^+^ patients with acute influenza infection and memory pools across different human tissues, highly relevant to the Indigenous population.

### Distinct pMHC tetramer staining patterns in HLA-A24 donors reflect KIR3DL1 binding

It was apparent from the tetramer-enrichment assays that some healthy donors contained large populations of HLA-A24-tetramer-binding CD8^+^ T-cells prior to enrichment (up to 6% of CD8^+^ T-cells) **(Fig. 7a)**. This appeared to be donor-dependent but not entirely CD8^+^ T-cell specificity-dependent. We found such oversized (0.32%-6.73% in unenriched PBMCs) tetramer^+^CD8^+^ T-cell populations for A/PB1_498_ in 10 out of 23 donors and in 14 out of 26 donors for B/NP_165_ tetramers, but not for B/NA_32_ (0/4 donors), which was further enriched with TAME (**Fig. 7a,b**). It is important to note that our tetramer analyses in Figure 5 excluded this oversized low intensity staining tetramer-binding CD8^+^ T-cell population. Such oversized tetramer-binding CD8^+^ T-cell population could potentially be a unique HLA-A24-tetramer binding phenomenon occurring in selected donors and hence potentially impair TCR-specific CD8^+^ T-cell binding. Therefore, we sought to better understand HLA-A24-tetramer binding in donors with conventional and largely oversized HLA-A24-tetramer CD8^+^ T-cell populations.

**Figure 7.**
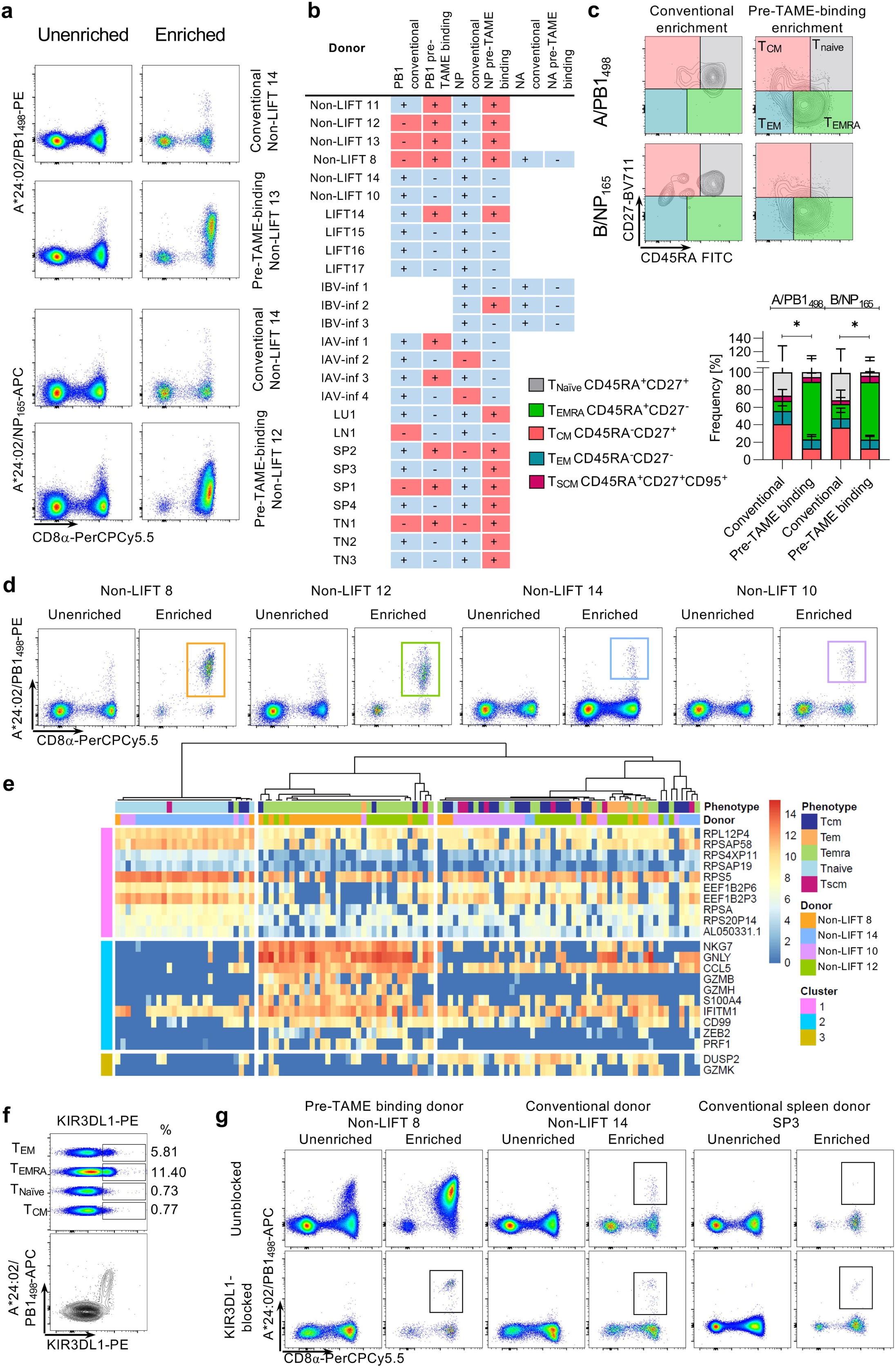
Comparison of conventional and KIR-binding CD8^+^ T-cells. **(a)** Staining pattern of donors with conventional-size tetramer staining and donors with oversized tetramer^+^CD8^+^ T cell populations for the A/PB1_498-505_ and B/NP_165-173_ tetramers pre- and post-enrichment. **(b)** Presence (+) or absence (-) of conventional (blue) and oversized tetramer^+^CD8^+^ T cell populations (red) populations across all samples tested. **(c)** Phenotypic analysis of tetramer-enriched CD8^+^ T-cells from conventional-size and oversized populations for both A/PB1_498-505_ and B/NP_165-173_ tetramers. **(d)** Tetramer staining pattern of conventional and oversized-binding staining for scRNAseq pre- and post-enrichment. **(e)** Unsupervised clustering of mRNA expression of enriched tetramer-binding CD8^+^ T-cells (n=30 cells per donor). All listed genes in the SC3 plot all genes listed have a p-value < 0.05 and AUC > 0.65. **(f)** KIR3DL1 expression on different CD8^+^ T-cell phenotypes in non-LIFT 8 (top panel) and co-staining of A/PB1_498-505_tetramer and KIR3DL1 without enrichment. **(g)** Effects of KIR3DL1 blocking on conventional and pre-TAME-binding tetramer^+^CD8^+^ T-cells pre- and post-enrichment.

Phenotypic analyses comparing tetramer-enriched fractions revealed that tetramer binding CD8^+^ T-cells of donors with oversized populations were predominantly of the CD45RA^+^CD27^-^ effector (T_EMRA_) phenotype (mean 73.3% and 71.7% for PB1_498_ and NP_165_, respectively), while those from donors with conventional tetramer^+^CD8^+^ T-cells were predominantly T_CM_ (mean 31.6 and 45.5%), T_EM_ (10.3 and 8.3%) and T_Naive_ (12.5 and 22.6%) in phenotype (**Fig. 7c**). To determine factors underlying this phenomenon, we performed scRNAseq on single-cell-sorted TAME-enriched A/PB1_498-505_^+^CD8^+^ T-cell populations from two donors with oversized populations (non-LIFT 8 and 12) and two donors with conventional-size populations (non-LIFT 14 and 10) (**Fig. 7d**). Unsupervised hierarchical clustering analysis revealed three gene clusters (**Fig. 7e**). Highly expressed genes from Cluster 1 were associated with A/PB1_498_^+^CD8^+^ T_Naïve_ cells predominantly from donor 14. Most notably, A/PB1_498_^+^CD8^+^ T_EMRA_ cells from donors 8 and 12 (oversized population) were grouped together and highly expressed genes from Cluster 2, characterized by high levels of T-cell effector genes NKG7, GNLY, CCL5 and granzymes B and H (GZMB and GZMH) but not K (GZMK), found in Cluster 3. In contrast, A/PB1_498-505_ ^+^CD8^+^ T_EMRA_ cells from donors 14 and 8 were grouped with highly expressed genes from Cluster 1 and 3, but not Cluster 2 except for CCL5 and IFITM1 genes, revealing distinct characteristics of A/PB1 ^+^CD8^+^ T_EMRA_ cells within the two tetramer-binding populations.

NKG7 (natural killer cell granule protein 7) and GNLY (granulysin) are key CD8^+^ T-cell effector genes^54^ located on the same immunoregion locus containing all the natural killer-receptor genes including the killer cell immunoglobulin-like receptors (KIR), within the leukocyte receptor complex (1Mb, chromosome 19q13.4)^55^. Since NKG7 and GNLY were the top-hit genes associated with A/PB1_498_^+^CD8^+^ T_EMRA_ cells from donors 8 and 12, we hypothesised that a KIR was interacting with the peptide/HLA-A24 complex. KIR are expressed by a proportion of CD8^+^ T-cells^56^ and KIR3DL1 in particular has been previously shown to bind some but not all A24 pMHC tetramers^57^ implying a degree of selectivity in the interaction. Staining for KIR3DL1 revealed its expression on CD27^-^CD8^+^ T-cells, with the highest frequency of KIR3DL1^+^ cells detected in the T_EMRA_ population in a donor that exhibited strong Pre-TAME tetramer binding (**Fig 7f**). Co-staining with the A/PB1_498-505_ tetramer showed that all tetramer-binding CD8^+^ T-cells were positive for KIR3DL1, indicating that KIR3DL1 could potentially be binding to the tetramers (**Fig 7f**). Blocking of KIR3DL1 prior to tetramer-staining markedly reduced the oversized population after TAME enrichment, to the levels of conventional tetramer^+^CD8^+^ T cell pools, revealing the true A24/PB1_498_-specific CD8^+^ T-cell population **(Fig. 7g)**. Thus, much of the oversized population comprises tetramer-binding KIR3DL1^+^CD8^+^ T-cells with other TCR specificities. Future studies are needed to understand whether KIR3DL1 binding of peptide-HLA-A24 complexes are competing with TCR interactions to mount robust peptide-HLA-A24-specific CD8^+^ T-cell responses, thus impacting on influenza-specific immunity in Indigenous and non-Indigenous HLA-A24-expressing people at risk of severe influenza disease.

## DISCUSSION

Indigenous populations worldwide are highly affected by pandemic and, to a lesser degree, seasonal influenza disease. In line with previous studies^58–61^, our results show a high frequency of HLA-A24 allele expression in Indigenous populations in the Pacific region, an allele identified as an influenza mortality-associated allomorph^30^. In our cohort of Indigenous Australians, 36% of individuals expressed at least one HLA-A24 allele. With little understood about the nature of the HLA-A24-restricted influenza-specific CD8^+^ T-cell response, there was a need to identify the breadth of influenza CD8^+^ T-cell epitopes for this at-risk population. Our analysis of previously published epitopes revealed a small number of HLA-A24-resticted IAV epitopes reported as immunogenic targets, while no IBV targets for HLA-A24 were known. Our in-depth mass-spectrometric approach defined the breadth of peptides presented by HLA-A*24:02 during IAV and IBV infection (**Tables 1 and 2**) and provided important insights into the characteristics of the associated CD8^+^ T-cell responses that could predispose to more severe influenza disease. Of the 52 peptides presented by HLA-A*24:02 during IAV infection, most mapped to PB2 (18 peptides, 35%) and PB1 (14 peptides, 27%) viral proteins, with no peptides originating from NA or M1. Consistent with this preference for PB2 and PB1 peptides, the CD8^+^ T-cell response in IAV-infected HHD-A24 mice focused mostly on four epitopes from PB1 (PB1_216-222_ and PB1_498-505_; 36% primary splenic response) and PB2 (PB2_549-557_ and PB2_549-559_; 31% primary splenic response). In HLA-A*24:02^+^ donors, memory responses to overlapping peptides PB1_496/498-505_ were consistently observed, while interesting differences where seen in the hierarchy of other epitope-specific responses, with Indigenous donors responding to PA_649-658_ and NP_39-47_, and non-Indigenous donors instead responding to the PB2_549-557_ peptide. Such differential response characteristics, possibly related to HLA co-expression or infection history, are important considerations for the design of T-cell vaccines for high risk Indigenous populations. Importantly, CD8^+^ T-cells specific for the dominant A/PB1_498-505_ peptide were identified with an activated phenotype in the blood of patients with acute IAV infection and across different human tissues, including a population of T_RM_ cells in the lung, providing evidence of their involvement in the influenza-specific response.

Hertz *et al*. previously showed that HLA-A24 has a low targeting efficiency for conserved regions of the pH1N1 virus, which was indicative of low cross-reactive memory responses that may have contributed to the impaired pH1N1 CD8^+^ T-cell immunity observed in HLA-A24^+^ individuals during the 2009 pandemic^30^. Our data reveal that the variable HA and NA viral glycoproteins play a minimal role in HLA-A24-restricted CD8^+^ T-cell immunity to IAV. Instead, the focus on epitopes from PB1 and PB2 that are well conserved across virus strains circulating in South-East Asia and Australia suggests that the prominent HLA-A24-restricted CD8^+^ T-cell responses are likely to confer broad cross-reactive immunity to IAV. This is of key importance as the current T-cell vaccines in clinical trials focus mainly on structural proteins like NP, M1 and M2, and would therefore not elicit cross-protective CD8^+^ T-cell responses in HLA-A24^+^ individuals at risk of severe influenza disease.

In contrast to IAV, the protein origins of IBV peptides presented by HLA-A24 differed greatly. From 41 IBV-derived peptides, the majority originated from NP (9, 22% of total), while 8 were from the HA and NA (total of 39% for surface glycoproteins). In terms of immunogenicity, our data from transgenic mice showed that the immunogenic HLA-A24-binding peptides were predominantly derived from NP (40% of response) and NA (40% of the response). More importantly, numbers of CD8^+^ T-cells directed towards our novel epitopes were preserved during secondary IBV challenge, indicating optimal memory establishment and recall, which contrasted with the situation for secondary IAV challenge. As in mice, HLA-A24-restricted influenza-specific CD8^+^ T-cell responses in Indigenous and non-Indigenous human donors were also targeted towards NP, with NP_165-173_ and NP_164-173_ being prominent CD8^+^ T-cell specificities alongside CD8^+^ T-cell epitopes derived from NA, HA, PB2 and PA (**Table 6**). The breadth of the HLA-A24-restricted IBV response highlights the power of identifying epitopes with our mass-spectrometric approach and might explain, at least partially, why Indigenous populations have not been reported to be at risk from severe IBV disease. As for IAV, IBV epitope-specific CD8^+^ T-cells were activated during acute IBV infection in HLA-A24^+^ individuals and were found distributed across tissues including the lung in non-infected individuals.

Broadly cross-reactive CD8^+^ T-cell responses that provide universal immunity across multiple strains or subtypes of influenza viruses have a crucial role in protection from severe influenza disease^27^. Here we demonstrate cross-reactive responses between IBV lineages for the B/NP_165-173_ peptide, as well as cross-reactive IAV/IBV responses between the A/PB2_549-557_ peptide and IBV PB2_550-558_ variants in HHD-A24 transgenic mice (data not shown) and humans. The A/PB2_549-557_ peptide is conserved between H3N2 and H1N1 IAVs^62^, and shares 55% amino acid identity with the cross-reactive IBV PB2_550-558_ variants. Structures of HLA-A24 with A/PB2_549-557_ and B/PB2_550-558_ showed that the antigen-binding cleft and both peptides adopted a similar conformation, providing a structural basis for T-cell cross-reactivity between these epitopes. Interestingly, IBV was more effective than IAV at expanding cross-reactive CD8^+^ T-cells, suggesting that infection history may play a role in determining patterns of cross-reactivity and that selection of peptide sequences that promote greatest cross-reactivity is a consideration for universal influenza T-cell vaccines.

Structural analysis of the overlapping peptides A/PB2_549-557/559_ and B/NP_164/165-173_ showed that despite the difference of two or one amino acids in length respectively, these peptides each adopted different conformations with HLA-A24 and are likely to induce distinct TCR repertoires. In the case of B/NP_164/165-173_, responses to both peptides are equivalently immunodominant in HLA-A24^+^ individuals, providing breadth to the overall CD8^+^ T-cell response. However, only the A/PB2_549-557_ epitope showed immunogenicity in HLA-A24^+^ individuals, with the instability and bulged conformation of the longer A/PB2_549-559_ epitope potentially proving challenging for TCR recognition. Such intricacies in epitope presentation and CD8^+^ T-cell recognition offer opportunities to either maximize or tailor responses through vaccination.

Our present study not only provides comprehensive data on generating CD8^+^ T-cell immunity against severe influenza disease in HLA-A24-expressing Indigenous and non-Indigenous people worldwide but also unravels three potential reasons why IAV-specific CD8^+^ T-cells in HLA-A24-expressing individuals might be perturbed: (i) *The antigenic origin of HLA-A24 IAV epitopes*. The majority (62%) of IAV peptides presented by HLA-A24 are derived from PB1 and PB2, which is in stark contrast to previous studies in humans^53,63,64,65^ and mice^66^ showing that immunodominant peptides for other HLAs are derived predominantly from NP, PA or M1. This is problematic for the current vaccine candidates in clinical trials which do not have PB1 or PB2 component^49,67,68^, and also raises the question of whether HLA-A24-restricted influenza-specific CD8^+^ T-cell responses are equivalently robust and protective compared to immunodominant responses restricted by other HLA. (ii) *Qualitative deficiencies in the HLA-A24 IAV-specific CD8*^*+*^ *T-cell response*. In HHD-A24 mice, the magnitude and breadth of IAV-specific CD8^+^ T-cell responses were greatly reduced during secondary IAV (but not IBV) challenge compared to primary infection, implicating possible defects at memory establishment or recall levels. (iii) *Non-epitope-specific binding of peptide-HLA-A24 complexes to KIR*. This can possibly limit TCR recognition and thus TCR-specific activation of influenza-specific CD8^+^ T-cells. The above observations provide a platform for further investigations to understand the mechanisms driving greater risk of severe influenza disease in HLA-A24^+^ individuals.

Our findings provide important insights into the design of new T-cell-targeted vaccines and immunotherapy protocols to reduce influenza disease mortality and morbidity in Indigenous people globally. Development of a vaccine that induces long-lasting broadly cross-reactive CD8^+^ T-cell immunity would provide at least some level of protection against distinct influenza variants, even strains with pandemic potential. Such a vaccine would minimise influenza-related deaths in global populations, especially high-risk groups, which includes HLA-A24-expressing Indigenous and non-Indigenous people. Our comprehensive analysis of peptide presentation and immunogenicity across mouse and human HLA-A24 models defines the candidate IBV and IAV peptides needed for a CD8^+^ T-cell-targeting vaccine that is effective in HLA-A24^+^ individuals. Understanding how best to augment these key responses to confer stronger protective immunity is the next step.

## Data Availability

All data are available upon request.

## ACKNOWLEDGEMENTS

HHD-A24 transgenic HHD mice were developed by Dr FrancCois Lemonnier (Pasteur Institute, Paris, France). We thank the Monash Macromolecular Crystallization Facility staff, and the staff at the Australian synchrotron for technical assistance. This work is supported by the National Health and Medical Research Council (NHMRC). This research was undertaken in part using the MX2 beamline at the Australian Synchrotron, part of ANSTO, and made use of the Australian Cancer Research Foundation (ACRF) detector.

## AUTHOR CONTRIBUTIONS

LH, PTI, EBC, THON, CES, LL, NAM, SG, JR and KK designed experiments. LH, PTI, EBC, THON, MK, CESS, NAM, AN, CS, HH, EJG, LL, BC and SG performed experiments. SR, TCK, ACC, MR, GPW, LMW, TL, SIM, ME, SGT, JD, AM, SYCT, PS, KF, DCJ, AB provided reagents and/or samples. LH, PTI, EBV, THON, NAM, AN, SR, FL, JR, SG, AWP and KK analyzed data. LH, THON, PTI, SG and KK wrote the manuscript. All authors read and approved the manuscript.

## LEGENDS TO SUPPLEMENTARY FIGURES

**Supplementary Figure 1. Further analysis of LC-MS/MS data sets. (a-c)** Peptide length distributions identified at a 5% FDR in each HLA isolation performed shown as a proportion of the peptides identified in the data set (n=number of peptides in the data set). For uninfected and HKx31 samples, data shown are based on searches against the human proteome, HKx31 proteome and 6 reading frame HKx31 genome translation. For B/Malaysia samples, data are based on searches against the human proteome, B/Malaysia proteome and 6 reading frame B/Malaysia genome translation. **(a)** Peptides from 12 data sets isolated from CIR.A24 using w6/32 (pan class I antibody) at different timepoints of infection with IAV or IBV (or uninfected). Asterisks represent data sets where DT9 was not used to deplete HLA-C*04:01 prior to w6/32 isolation and may contain increased levels of HLA-C*04:01 ligands. All data sets contain low levels of peptides presented by HLA-B*35:03 of CIR. **(b)** 3 data sets containing peptides isolated from the endogenous HLA class I of the CIR cell line, either through isolation from non-transfected CIR using w6/32 (CIR 16hrs HKx31 (w6/32)), hence containing HLA-C*04:01 and HLA-B*35:03, or through specific isolation of HLA-C*04:01 from CIR.A24 using the DT9 antibody. **(c)** HLA class II peptide ligands from 2 data sets isolated from CIR.A24 cell line using LB3.1 (HLA-DR), SPV-L3 (HLA-DQ), and B721 (HLA-DP) antibodies. **(d)** The sequence logo generated from human-derived 9mer peptides isolated in (b) (non-redundant by sequence, 5% FDR), and filtered for peptides identified in HLA class II isolations (c). **(e,f)** Pie charts showing the distribution of IAV (HKx31) derived peptides across the viral proteome potentially bound to HLA-B*35:03 and HLA-C*04:01 (e) and HLA-II of CIR cells (f). Details of HLA isolation antibodies, influenza peptide sequences, their confidence of sequence assignment, and potential binding assignment to the HLA molecules expressed by CIR.A24 are provided in **Supplementary Data 1**.

**Supplementary Figure 2. Comparison of epitope-specific CD8^+^ T-cells in primary and secondary IAV and IBV infection**. Total number of epitope-specific CD8^+^ T-cells calculated by the frequency of IFNγ^+^CD8^+^ T-cells in the INF-γ ICS assay multiplied by the total number of splenocytes per spleen.

**Supplementary Figure 3. Crystal structures of HLA-A*24:02 presenting the PB2**_**549-559**_ **peptide. (a)** Depiction of the PB2_549-559_ peptide presented by HLA-A*24:02. **(b)** Overlay of PB2_549-557_ (red) and PB2_549-559_ (maroon) peptides presented by HLA-A*24:02. The two extra residues of the PB2_549-559_ peptide are shown in yellow. **(c)** The P9-Trp of the PB2_549-557_ peptide (red) is forming a network of interactions with Tyr at positions 116, 118 and 123 and Leu95 within the peptide-binding cleft. **(d)** Lys146 of the α2-helix of HLA-A*24:02 also interacts with the carboxylic group of the PΩ residue of the PB2_549-557_ 9mer peptide (red) but **(e)** faces outside of the cleft and does not interact with the P9-Trp of the PB2_549-559_ 11mer peptide (maroon).

**Supplementary Figure 4. Crystal structures of HLA-A*24:02 presenting NP**_**164-173**_ **and NP**_**165-173**_ **peptides. (a-c)** Structures of NP_165-173_ (**a**, blue stick), NP_164-173_ (**b**, light blue stick) and **(c)** an overlay of NP_165-173_ (blue) and NP_164-173_ (light blue) presented on the HLA-A*24:02 molecule. **(d)** N-terminal side view overlay of NP_165-173_ (blue) and NP_164-173_ (light blue) peptides showing the largest structural difference at the Cα atom of the P6/7-Arg shown by the red dashed line betweenNP_165-173_ (blue) and NP_164-173_ (light blue), respectively. **(e)** The P5-Ile and P7-Val of the NP_165-173_ 9mer peptide (blue) are half-buried between the peptide backbone and the HLA α2-helix, and together with the P4-Pro, form a hydrophobic patch at the centre of the peptide. **(f)** The substitution of P2-Tyr (NP_164-173_) (light blue) for P2-Phe (NP_165-173_) (blue) occurs without major structural rearrangement of the B pocket residues (white sticks), as both residues are large and aromatic. **(g)** The additional residue of NP_164-173_ (light blue) changes the secondary anchor residue at P3 of the peptides from a small P3-Ser (NP_165-173_) to a large P3-Phe (NP_164-173_).

**Supplementary Figure 5. Electron density maps of the peptide-HLA-A*24:02 structures**.

Peptide presentation in the antigen binding cleft of HLA-A*24:02 showing electron density omit maps (Fo-Fc) at 3.0σ (green) and electron density maps (2Fo-Fc) at 1.0σ (blue), for **a-d** and **i** and **e-h** and **j** respectively. The HLA-A*24:02 is represented as white cartoon, while the peptides are represented in sticks and coloured in red (PB2_549-557_) pink (PB2_549-557B_), maroon (PB2_549-559_), blue (NP_165-173_), and light blue (NP_164-173_).

**Supplementary Data 1. Peptides identified from IAV (HKx31) and IBV (B/Malaysia)** Sequences of IAV (HKx31) and IBV (B/Malaysia)-derived peptides identified by LC-MS/MS analysis of the HLA class I and II immunopeptidomes of CIR and CIR.A24. For each peptide, the modifications with which it was identified, source protein, start site within the source protein, and the confidence of assignment for the data sets within which it was identified, are noted. For each data set, the cell line, infecting virus, antibody used (and any antibody depletion prior), time of infection and confidence cut-off for a 5% FDR are shown. Identifications above this confidence in each data set are in bold, those below are in italics and have increased likelihood of being false positive identifications. Naming of data sets match those in Supplementary Fig. 1. The predicted binding affinities (nM) and %rank for HLA-A*24:02, HLA-B*35:03 and HLA-C*04:01 for all 8-14mer peptides as calculated by NetMHCpan4.0 are shown. For HKx31, data sets derived from the sequential isolation of HLA from the same sample are noted. For B/Malaysia, previous identification in HLA isolations from B/Malaysia infected CIR and CIR.A*02:01 in Koutsakos *et al*.^27^ are also noted. Colour fill represents isolations with w632 (blue), DT9 (yellow) and mixed class II antibodies (green). The “Best Explanation” column denotes the HLA hypothesised to present a given peptide based on appearance across the data sets and predicted binding by NetMHCpan4.0.

## METHODS

### Human blood and tissue samples

Human experimental work was conducted according to the Declaration of Helsinki Principles and according to the Australian National Health and Medical Research Council Code of Practice. All blood and tonsil donors provided written consent prior to study participation. Lung tissues, lymph node and spleen samples were obtained from deceased organ donors after receiving written informed consent from next-of-kin. Lungs were sourced from the Alfred Hospital’s Lung Tissue Biobank. Lymph node and spleen were provided by DonateLife Victoria. Buffy packs were sourced from the Australian Red Cross Lifeblood (West Melbourne, Australia). Human experimental work was approved by the University of Melbourne Human Ethics Committee (ID 1955465.2, 1443389.4, 1441452.1 and 1954302.1), the Australian Red Cross Lifeblood Ethics Committee (ID 2015#8), the Alfred Hospital Ethics Committee (ID 280/14), Monash Health Human Research Ethics Committee (HREC) (ID HREC/15/MonH/64, RMH local reference number 2016/196), HREC of Northern Territory Department of Health and Menzies School of Health Research (ID 2012-1928) and Tasmanian Health and Medical HREC (ID H0017479). Human PBMCs and cells from tissues were isolated and cryopreserved as previously described^70^. Indigenous donors (LIFT) were recruited as described before^29^.

### HLA typing and analysis of human PBMCs

NGS HLA typing for HLA class I and class II on genomic DNA isolated from granulocytes was performed by the Victorian Transplant and Immunogenetics Service (West Melbourne, VIC, Australia). Co-expression Circos plots were generated using R V.4.0 (R Core Team, Vienna, Austria), RStudio: Integrated Development for R (RStudio, Inc., Boston, USA) and the cyclize package^71^.

### Cell lines, viruses and peptides

Class I-reduced (CIR) B-LCL express low levels of HLA-A and B, but normal levels HLA-C*04:01^36,72^. CIR.A24 cells were generated by transfecting CIR cells with HLA-A*24:02 in the pcDNA3.1(+)-hygro vector. Cells were maintained in RF10 medium (RPMI-1640 with 10% heat-inactivated FCS, 100mM MEM non-essential amino acids, 55 mM 2-mercaptoethanol, 5 mM HEPES buffer solution, 1mM MEM sodium pyruvate, 1 mM L-glutamine, 100 U ml^-1^ penicillin and 100 mg ml^-1^ streptomycin (purchased from Gibco/Thermo Fisher Scientific) with the addition of 0.3 mg/mL hygromycin-B (Life Technologies) for CIR.A24 cells. Influenza A (A/HKx31 & A/Puerto Rico/8/1934 (A/PR8)) and B viruses (B/Malaysia/2506/04 & B/Phuket/3073/2013) were grown for 3 days at 35°C in the allantoic cavity of 10 day-old embryonated chicken eggs. Viral titres were determined performing semisolid overlay plaque assay on MDCK cells (ATCC). Influenza B viruses were kindly provided by Steve Rockman (Seqirus, Australia). Influenza peptides were synthesized by GenScript (Piscataway, NJ, USA) with a purity >80% and reconstituted at 1mM in 100% DMSO. Cell lines tested negative for mycoplasma by PCR using primers 5’-YGCCTGVGTAGTAYRYWCGC-3’ and 5’-GCGGTGTGTACAARMCCCGA-3’.

### Expansion of antigen-specific memory CD8^+^ T-cells from human PBMC

Cryopreserved PBMCs (3.3-5×10^6^) from healthy non-LIFT and LIFT donors were used to expand antigen-specific CD8^+^ T-cells modified from Koutsakos *et al*^27^. In brief, one-third of PBMCs were pulsed with a pool of up to 31 peptides (including circulating variants, **Supplementary Table 4**) at a total concentration of 10µM at 37°C in RPMI. After 1 hr, cells were washed twice with RPMI and mixed with the remaining autologous PBMCs. To expand virus-specific CD8^+^ T-cells, infected CIR.A24 cells were washed twice with serum-free RPMI to remove excess FCS and infected with A/HKx31 or B/Malaysia/2560/2004 at a MOI of 5 and incubated at 37□. After 1 hr, RF10 was added and cells were incubated for further 11 hrs at 37□ before cells were placed at 4□ for 14 hrs. Infected CIR.A24 cells were washed twice to remove any residual virus and added at a 1:10 ratio to PBMCs. For additional stimulation, virus-expanded PBMCs were restimulated by addition of virus-infected CIR.A24 cells on day 8 at a 1:10 ratio. Cells were then incubated for a total 10-15 days in RF10 media with 10U/ml of recombinant human IL-2 (Roche Diagnostics, Mannheim, Germany) being added on day 4 and half-media changes every 1-2 days onwards.

### T-cell restimulation and intracellular cytokine staining

To identify epitope-specific CD8^+^ T-cells that expanded after stimulation, cells from day 10-15 cultures (2×10^5^ cells/well) of peptide-expanded PBMCs were mixed with peptide-pulsed CIR.A24 at a 1:3 ratio while virus-expanded PBMCs were restimulated by direct peptide addition (1 μM). Cells were incubated for 5 hrs in the presence of Brefeldin A (BD Golgi Plug), Monensin (BD Golgi Stop) and anti-CD107a FITC at 37□. Cells were stained with panel 2 (**Supplementary Table 6**) and analyzed using flow cytometry (BD Fortessa) and FlowJo v10 (BD).

### Large-scale infection for immunopeptidome analysis

For large scale infections, CIR or CIR.A24 were cultured to high density in RF10 media slowly rotating in 17dm^2^ filter-capped roller bottles (Corning) at 37°C, 5% CO_2_. Cells were harvested and infected with influenza A or B virus at a MOI of 5 in RPMI at a density of 1 × 10^7^ cells/mL in 50mL tubes for 1 hour at 37°C with slow rotation. Infected cells were returned to roller bottles with the addition of 1:1 conditioned media:RF10 to a final density of 1.4 × 10^6^ cells/mL and incubated a further 1-15 hours (37°C, 5% CO_2_, slow rotation). HLA expression and infection efficacy were validated by surface staining ∼10^6^ cells with anti-HLA class I PE-Cy7 (1:200 in PBS; Biolegend, Cat# 311430; 30min, 4°C), prior to washing in PBS, fixation in 1% paraformaldehyde (ProSciTech) in PBS (20min, room temperature), and intracellular staining with anti-NP FITC for influenza A (Clone 1331, GeneTex Cat# GTX36902) or influenza B (Clone H89B, ThermoFisher Cat# MA1-7306) (1:200 in 0.3% saponin [Sigma] in PBS, 30min, 4°C). Cells were washed in PBS and acquired by flow cytometry using a BD LSRII flow cytometer running BD FACSDiva software, and analysed using FlowJo_v10 (BD). Remaining cells were harvested by centrifugation in 500mL V-bottom flasks (3283*g*, 15min, 4°C), washed in PBS, snap frozen as cell pellets in liquid nitrogen, and stored at -80°C until use. Uninfected cells were harvested and snap frozen as for infected cells.

### Liquid Chromatography-tandem mass spectrometry (LC-MS/MS) analysis of HLA-bound peptides

Cell pellets of 0.7-1.3×10^9^ CIR or CIR.A24 were lysed via cryogenic milling (Retsch Mixer Mill MM 400), resuspension in 0.5% IGEPAL CA-630, 50 mM Tris-HCl pH8.0, 150 mM NaCl and protease inhibitors (cOmplete Protease Inhibitor Cocktail Tablet; Roche Molecular Biochemicals) and incubation at 4°C for 1 hour with slow rotation. Lysates were cleared by ultracentrifugation and HLA isolated by immunoaffinity purification using protein-A-sepharose-bound antibodies as described^38,73^. Antibodies were either w6/32 (pan class I) alone or sequential DT9 (HLA-C specific), w6/32 (pan class I) and mixed class II (equal amounts LB3.1, SPV-L3 and B721, capturing HLA-DR, -DQ and -DP, respectively).

Peptide/MHC complexes were dissociated, and fractionated by reversed phase high performance liquid chromatography (RP-HPLC) as described^27,38,74^. 500µL fractions were collected throughout the gradient, and the peptide containing fractions combined into 9 pools, vacuum-concentrated and reconstituted in 15µL 0.1% formic acid (Honeywell) in Optima™ LC-MS water. Reconstituted fraction pools were analysed by LC-MS/MS using a SCIEX 5600+ TripleTOF mass spectrometer equipped with a Nanospray III ion source as previously described^74^.

### LC-MS/MS data analysis

Spectra were searched against a proteome database consisting of the human proteome (UniProt/Swiss-Prot v2016_04), and either the A/X31 or the B/Malaysia/2506/2004 proteome plus a 6 reading frame translation of the viral genome, using ProteinPilot software (version 5.0, SCIEX), considering biological modifications and employing a decoy database to calculate the false discovery rate (FDR). Subsequent analyses were based on the best hypothesis for distinct peptides. Sequence motifs were generated utilizing peptides assigned at confidences greater than that required for a 5% FDR using Seq2logo2.0^69^ (default settings). Likely HLA-A*24:02 binders were determined based on appearance across the experiments/antibodies and predicted binding (netMHCpan4.0). For peptides identified in their native form (and lacking Cys residues) that were synthesised for functional analysis, fragmentation patterns and retention times of representative spectra were compared to the synthetic and the quality of the match described (**Supplementary Data 1**).

### HLA-A*24:02 HHD mouse studies

All mouse studies were overseen by the University of Melbourne Ethics Committee (#171408). HHD-A24 mice were generated by François Lemonnier as described previously^42^. These mice express a chimeric MHC-I that consists of the murine α3 and transmembrane domain and the human α1 and α2 domain covalently linked the human β2m. Mouse infections were performed as described previously^24^. In brief, 6-12 week-old mice were infected intranasally with 30 µl of either 100 pfu of A/X-31 or 200 pfu of B/Malaysia/2506/2004 in PBS under isoflurane anaesthesia. For secondary challenge, mice were infected 6-8 weeks after primary infection with 200 pfu of A/PR8 or 400 pfu B/Phuket/3073/2013. To identify immunogenic peptides, spleen and bronchioalveolar lavage (BAL) were isolated on day 10 or day 8 for secondary infection, respectively. Spleen single cell suspensions were prepared and incubated for 1 h at 37□ in Affinipure Goat anti-mouse IgG+IgM (Jackson Immunoresearch)-coated panning plates to deplete B cell populations. BAL was combined from 3-5 mice to achieve sufficient T-cell numbers. Single cell suspensions were then stimulated with peptide pools or single peptides at 1 µM in the presence of Brefeldin A (BD Golgi plug) for 5h at 37□ in RF10 with 10 U/ml IL-2 followed by staining with panel 1 (**Supplementary Table 6**). Cells were analyzed using BD Fortessa and FlowJo v10 (BD). For immunization studies, mice were vaccinated with 30nmol of NA_32-40_, NP_392-400_ and NP_165-173_ emulsified in complete (Prime) or incomplete (Boost) Freund’s adjuvant. 50μl vaccine was injected on both sides at the base of the tail. Control mice were injected emulsified adjuvant without peptides. Two weeks after prime, mice were boosted and challenged with 1×10^3^ pfu B/Malaysia/2506/2004 intranasally 7 days after boost. On day 6 and 7 after infection lungs were isolated to determine viral load with a plaque assay as described before^27^. Cytokines in the bronchoalveolar lavage were assessed with the BD cytometric bead array kit as described elsewhere^27^.

### Protein expression, purification and crystallization

Soluble HLA-A*24:02 heterodimers containing either A/PB2_549-557_, A/PB2_549-559_, B/PB2_549-557_, B/NP_165-173_, or B/NP_164-173_ peptide were prepared as previously described^53^. In brief, a truncated HLA-A*24:02 construct encompassing the extracellular part of the HLA molecule (residues 1-276), and human beta-microglobulin (β2m) were expressed separately in a BL21-pLyS *Escherichia coli* strain as inclusion bodies. The inclusion bodies were subsequently extracted, washed and resuspended into a solution containing 6M guanidine. Each pHLA complex was then refolded into a cold refolding solution (100 mM Tris-HCl pH 8, 2 mM Na-EDTA, 400 mM L-arginine-HCl, 0.5 mM oxidized glutathione, 5 mM reduced glutathione) by adding 30mg of HLA heavy chain, 20mg of β2m and 4mg of peptide. The refolding solution was then dialyzed in 10mM Tris-HCl pH 8, and the protein was purified by a succession of affinity column chromatography.

### Crystallization, data collection and structure determination

Crystals of the pHLA-A*24:02 complexes were grown by the hanging-drop, vapour-diffusion method at 20°C with a protein/reservoir drop ratio of 1:1 with seeding at a concentration of 6 mg/mL in the following with conditions: A/PB2_549-557_: 20% PEG3350, 0.2M Na acetate ; PB2_549-559_: 24% PEG8K, 0.1 HEPES pH 7.5, 5% v/v Ethyl acetate; B/PB2_549-557_: 24% PEG8K, 0.1 HEPES pH 7.5, 2% isopropanol, 5% w/v PolyvinylpyrrolidoneK15 ; B/NP_165-173_: 19% 3350, 0.2 MgCl_2_; NP_164-173_: 20% PEG8K, 0.2 MgCl_2_, 0.1 Tris-HCl pH 8.5. The crystals were soaked in a cryoprotectant solution containing mother liquor solution with the PEG concentration increased to 30% (w/v) and then flash frozen in liquid nitrogen. The data were collected on the MX1 and MX2 beamlines^75^. Manual model building was conducted using the Coot software^76^ followed by maximum-likelihood refinement with the Buster program^77^. The final models were validated using the Protein Data Base validation web site and the final refinement statistics are summarized in **Supplementary Table 5**. All molecular graphics representations were created using PyMol^78^.

### Thermal stability assay

Thermal shift assays were performed to determine the stability of each pHLA-A*24:02 complex using fluorescent dye Sypro orange to monitor protein unfolding. The thermal stability assay was performed in the Real Time Detection system (Corbett RotorGene 3000), originally designed for PCR. Each pHLA complex was in 10 mM Tris-HCl pH8, 150 mM NaCl, at two concentrations (5 and 10 mM) in duplicate, was heated from 25 to 95°C with a heating rate of 1°C/min. The fluorescence intensity was measured with excitation at 530 nm and emission at 555 nm. The Tm, or thermal melt point, represents the temperature for which 50% of the protein is unfolded.

### Tetramer-associated magnetic enrichment in humans

TAME was performed on PBMCs (7.5×10^6^-2.7×10^8^) of healthy, IAV- or IBV-infected donors, as well as lymphocytes isolated from tonsils, lung and pancreatic lymph nodes (panLN) to detect CD8^+^ T-cells specific for IAV and IBV as described previously^25,27^. PMHC-I monomers were made in-house^79^ and conjugated at an 8:1 molar ratio to PE- or APC-labelled streptavidin (SA) to generate tetramers. Cells were FcR-blocked and stained with APC or PE conjugated tetramers at a 1:100 dilution for 1h at RT, washed twice then incubated with anti-PE and anti-APC MicroBeads (Milenty Biotec). Unenriched, flow-through and enriched fractions were surface stained with panel 3 (PBMC) or 4 (SLO and lung) (**Supplementary Table 6**). After 30min staining on ice, cells were fixed for 20 min in 1% PFA and acquired by flow cytometry. In some experiments, KIR3DL1 blocking was achieved by the addition of anti-human NKB1 antibody (DX9, Cat 555964, BD Pharmingen) at 1:100 during the FcR-blocking step.

### Single-cell mRNAseq

A/PB1_498-505_^+^ CD8^+^ T-cells were single cell sorted into 96 well plates containing lysis buffer (1µl RNase inhibitor and 19µl Triton X-100) after TAME on a BD Aria III sorter. Libraries were generated as described previously^27^. A Nextera XT DNA Library Prep Kit was used for the generation of sequencing libraries and sequencing performed on a NextSeq500 platform with 150-base par high-output paired-end chemistry for 30 tetramer^+^ cells/donor (120 cells total).

### Bioinformatical analysis

Gene expression was analysed as previously described^27^. Briefly, quality of scRNA-seq was assessed with FastQC. TopHat2 with default parameters was used to align sequences to the Ensembl GRCh38 reference genome. A total of 112 out of 120 analysed cells passed quality control and was used for further analysis. Gene expression was quantified utilizing Cufflinks suit (v 2.2.1) where FPKM was assessed using CuffQuant and values normalized based on total mRNA content with CuffNorm. Clustering was performed utilizing SC3^80^. Pheatmap in R was used to visualize Heatmaps.

### Statistical analyses

Statistical analysis was performed using Graphpad Prism (v8.4.2, Graphpad, USA). Non-parametric unpaired data was analysed performing Mann-Whiteney test whereas paired data was analysed with Wilcoxon matched-pairs signed rank test. Two-tailed analysis was performed with p<0.05 (*) or p<0.01 (**) indicated.

### Data availability statement

The mass spectrometry proteomics data have been deposited to the ProteomeXchange Consortium via the PRIDE [1] partner repository with the dataset identifier PXD020292. Username; Password: 3dTWu8SU

