## Supplemental Tables for "CD8^+^ T-cell landscape in Indigenous and non-Indigenous people restricted by influenza mortality-associated HLA-A*24:02 allomorph"

**Table 1.** Human immunodominant A24 epitopes

| Peptide | Sequence | Virus origin | Source |
| --- | --- | --- | --- |
| PA_649-658_ | LYASPQLEGF | IAV | Previous identified/Mass-spectrometry |
| PB1_496-505_ | FYRYGFVANF | IAV | Previously identified |
| PB1_498-505_ | RYGFVANF | IAV | Previous identified/Mass-spectrometry |
| PB2_549-557_ | TYQWIIRNW | IAV | Previous identified/Mass-spectrometry |
| PA_457-465_ | KYVLFHTSL | IBV | Mass-spectrometry |
| PB1_503-511_ | NFAMELPSF | IBV | Mass-spectrometry |
| PB2_550-558_ | TYQWVLKNL | IBV | Mass-spectrometry |
| HA_552-560_ | YYSTAASSL | IBV | Mass-spectrometry |
| NA_32-40_ | LYSDILLKF | IBV | Mass-spectrometry |
| NP_165-173_ | YFSPIRVTF | IBV | Mass-spectrometry |

**Supplementary Table 1.** Donor demographics

| Donor ID | Age | Sample | HLA-A | HLA-B | HLA-C | HLA-DRB1 | HLA-DPB1 | HLA-DQB1 | IAV peptide expansion | IBV peptide expansion | IBV virus expansion | IAV virus expansion | TAME |
| --- | --- | --- | --- | --- | --- | --- | --- | --- | --- | --- | --- | --- | --- |
| LIFT1 | 43 years | PBMC | 24:02, 34:01 | 15:21, 56:01 | 01:02, 04:03 | 04:12, 14:08 | 05:01, 16:01 | 04:02, 05:03 | X |  |  |  |  |
| LIFT2 | 46 years | PBMC | 24:02 | 13:01, 56:01 | 01:02, 04:01 | 08:03, 14:02 | 04:01, 05:01 | 03:01, 06:01 |  | X |  |  |  |
| LIFT3 | 65 years | PBMC | 24:02 | 13:01, 40:02 | 01:02, 04:01 | 08:03 | 02:01, 04:01 | 06:01 | X |  |  |  |  |
| LIFT4 | 68 years | PBMC | 11:01, 24:02 | 39:01, 40:10 | 04:03, 12:03 | 08:03 | 02:01, 05:01 | 06:01 | X |  |  |  |  |
| LIFT5 | 39 years | PBMC | 02:01, 24:02 | 15:21, 56:01 | 04:03, 07:02 | 04:05, 14:08 | 05:01, 14:01 | 04:01, 05:03 |  | X |  |  |  |
| LIFT6 | 34 years | PBMC | 02:01, 24:02 | 07:02, 40:06 | 03:04, 07:02 | 11:01, 15:01 | 05:01 | 03:01, 06:02 | X |  |  |  |  |
| LIFT7 | 50 years | PBMC | 01:01, 24:02 | 08:01, 15:21 | 04:03, 07:01 | 03:01, 14:07 | 14:01, 16:01 | 02:01, 05:03 |  | X |  |  |  |
| LIFT8 | 34 years | PBMC | 24:02 | 40:01, 56:02 | 01:02, 03:03 | 08:03, 15:02 | 05:01 | 06:01 |  | X |  |  |  |
| LIFT9 | 21 years | PBMC | 24:02, 34:01 | 07:02, 56:01 | 01:02, 07:02 | 04:12, 15:01 | 02:01, 04:01 | 04:02, 06:02 |  | X |  |  |  |
| LIFT10 | 36 years | PBMC | 24:02 | 40:01, 40:02 | 01:02, 03:03 | 08:03, 15:02 | 02:01, 05:01 | 06:01 | X |  |  |  |  |
| LIFT11 | 24 years | PBMC | 24:02, 34:01 | 13:01 | 04:01, 15:02 | 04:12, 12:01 | 05:01 | 03:01, 04:02 |  | X |  |  |  |
| LIFT12 | 25 years | PBMC | 11:01, 24:02 | 13:01, 40:02 | 04:01, 15:02 | 04:05, 08:03 | 02:01, 04:01 | 03:01, 06:01 |  | X |  |  |  |
| LIFT13 | 30 years | PBMC | 24:02 | 13:01, 56:01 | 01:02,03:03 | 14:08 | 04:01 | 05:03 |  | X |  |  |  |
| LIFT14 | 21 years | PBMC | 24:02, 24:03 | 40:01, 52:01 | 03:03, 12:02 | 04:12, 15:02 | 04:01, 05:01 | 04:02, 06:01 |  |  |  |  | X |
| LIFT15 | 27 years | PBMC | 24:02 | 40:01, 56:02 | 01:02, 03:03 | 08:03 | 02:01 | 05:03, 06:01 |  |  |  |  | X |
| LIFT16 | 27 years | PBMC | 01:01, 24:02 | 56:01, 57:01 | 01:02, 06:02 | 07:01,15:02 | 04:01, 05:01 | 03:01, 03:02 |  |  |  |  | X |
| LIFT17 | 21 years | PBMC | 24:02, 24:06 | 40:01, 56:01 | 01:02, 03:03 | 04:12, 14:34 | 05:01 | 04:02, 05:03 |  |  |  |  | X |
| Non-LIFT1 | 29 years | PBMC | 01:01, 24:02 | 08:01, 14:02 | n.D. | n.D. | n.D. | n.D. | X |  |  |  |  |
| Non-LIFT2 | 30 years | PBMC | 03:01, 24:02 | 35:03, 44:02 | n.D. | n.D. | n.D. | n.D. | X |  |  |  |  |
| Non-LIFT3 | n.D. | PBMC | 02:07, 24:02 | 46:01, 58:01 | n.D. | n.D. | n.D. | n.D. | X |  |  |  |  |
| Non-LIFT4 | n.D. | PBMC | 03:01, 24:02 | 15:01, 47:01 | n.D. | n.D. | n.D. | n.D. | X |  |  |  |  |
| Non-LIFT5 | n.D. | PBMC | 24:02 | 40:02, 51:01 | n.D. | n.D. | n.D. | n.D. | X |  |  |  |  |
| Non-LIFT6 | 59 years | PBMC | 03:01, 24:02 | 15:01, 38:01 | 03:03, 12:03 | 13:01 | n.D. | n.D |  | X | X | X |  |
| Non-LIFT7 | 60 years | PBMC | 02:01, 24:02 | 15:01, 35:02 | 01:02, 04:01 | 08:01, 11:04 | n.D. | n.D. |  | X | X | X |  |
| Non-LIFT8 | 59 years | PBMC | 24:02 | 07:02 | 07:02 | 01:01, 10:01 | n.D. | n.D |  | X | X | X | X |
| Non-LIFT9 | 29 years | PBMC | 01:01, 24:02 | 35:01, 57:01 | 04:01, 06:02 | 01:01, 14:54 | n.D. | n.D |  | X | X | X |  |
| Non-LIFT10 | 38 years | PBMC | 02:07, 24:02 | 46:01, 67:01 | 01:02, 07:02 | 08:03, 09:01 | n.D. | n.D |  | X | X | X | X |
| Non-LIFT11 | 60 years | PBMC | 24:02, 25:01 | 07:02, 18:01 | n.D. | n.D. | n.D. | n.D. | X |  |  |  | X |
| Non-LIFT12 | 24 years | PBMC | 02:03, 24:02 | 27:06, 40:01 | 03:04, 04:01 | n.D. | n.D. | n.D. |  |  |  |  | X |
| Non-LIFT13 | 36 years | PBMC | 01:01, 24:02 | 08:01, 40:01 | 03:04, 07:01 | 04:04/23, 11:01 | n.D. | n.D. |  |  |  |  | X |
| Non-LIFT 14 | 34 years | PBMC | 01:01, 24:02 | 44:02, 57:01 | 06:02, 16:04 | 04:02, 07:01 | n.D. | n.D. |  |  |  |  | X |
| IAV-Inf 1 | 44 years | PBMC | 01:01, 24:02 | 08:01, 57:01 | 06:02, 07:01 | 03:01, 07:01 | 01:01, 04:01 | 02:01, 03:03 |  |  |  |  | X |
| IAV-Inf 2 | 86 years | PBMC | 24:02, 29:02 | 18:01, 51:01 | 07:01, 16:02 | 04:03, 15:01 | 01:01, 04:01 | 03:02, 06:02 |  |  |  |  | X |
| IAV-Inf 3 | 33 years | PBMC | 24:02, 32:01 | 07:02, 56:01 | 01:02, 07:02 | 01:01 | 04:01; 06:01 | 05:01 |  |  |  |  | X |
| IAV-Inf 4 | 77 years | PBMC | 24:02, 74:01 | 07:02, 51:01 | 01:02, 15:02 | 04:03, 15:01 | 04:01 | 03:02, 06:02 |  |  |  |  | X |
| IBV-Inf 1 | 52 years | PBMC | 11:01, 24:02 | 08:01, 15:01 | 03:03, 07:01 | 04:01, 15:01 | 03:01 | 04:02 |  |  |  |  | X |
| IBV-Inf 2 | 68 years | PBMC | 24:02, 24:03 | 40:06, 51:06 | 12:04, 15:07 | 04:03, 07:01 | 01:01, 02:01 | 03:02, 03:03 |  |  |  |  | X |
| IBV-Inf 3 | 63 years | PBMC | 03:01, 24:02 | 07:02, 51:01 | 0x:03, 07:02 | 09:01, 15:01 | n.D. | 03:03, 06:02 |  |  |  |  | X |
| Sp1 | 55 years | Spleen | 02:01, 24:02 | 27:05, 35:01 | 02:01, 04:01 | 01:01, 12:01 | n.D. | n.D. |  |  |  |  | X |
| Sp2 | 48 years | Spleen | 02:01, 24:02 | 07:02, 15:01 | 03:01, 07:01 | 13:01, 15:01 | n.D. | n.D. |  |  |  |  | X |
| Sp3 | 58 years | Spleen | 02:01, 24:02 | 27:05, 35:01 | 02:01, 04:01 | 01:01, 11:01 | n.D. | n.D. |  |  |  |  | X |
| Sp4 | 63 years | Spleen | 02:01, 24:02 | 07:02, 15:01 | 03:01, 07:01 | 13:01, 15:01 | n.D. | n.D. |  |  |  |  | X |
| LN1 | 55 years | Pancreatic Lymphnode | 02:01, 24:02 | 27:05, 35:01 | 02:01, 04:01 | 01:01, 12:01 | n.D. | n.D. |  |  |  |  | X |
| TN1 | 7 years | Tonsil | 24:02, 29:02 | 27:05, 42:01 | 02:02, 17:01 | 04;01, 08:04 | 02:01 | 03:01, 03:02 |  |  |  |  | X |
| TN2 | 42 years | Tonsil | 24:02, 32:01 | 15:01, 44:02 | 03:03, 05:01 | 04:01, 15:01 | 04:01, 05:01 | 03:02, 06:02 |  |  |  |  | X |
| TN3 | 4 years | Tonsil | 24:02, 29:02 | 51:01, 58:01 | 03:03, 07:18 | 08:04, 15:01 | 04:01 | 04:02, 06:02 |  |  |  |  | X |
| L1 | 41 years | Lung | 3, 24 | 15, 55 | 3, 3 | n.D | n.D | n.D |  |  |  |  | X |

n.D. = not determined

**Supplementary Table 2.** Influenza A peptides identified by immunopeptidomics.

| Peptide | Sequence | Virus origin | Source | Peptide Pool | Conservation^1^ | Predicted binding affinity (nM)^2^ | Comb. CTL epitope score^3^ | Immunogenic in human donors (of tested) | Immunogenic in mice (of tested) |
| --- | --- | --- | --- | --- | --- | --- | --- | --- | --- |
| PA_47-53_ | MYSDFHF | IAV | MS | 5 | 1 |  |  |  | 0/10 |
| PA_47-60_ | MYSDFHFINEQGES | IAV | MS | 5 | 0.52 | 18148 |  |  | 0/10 |
| PA_48-53_ | YSDFHF | IAV | MS | 5 | 1 |  |  |  | 0/10 |
| PA_130-138_ | YYLEKANKI | IAV | MS/iedb | 1 | 1 | 29 |  | 1/10 | 1/10 |
| PA_457-465_ | EYIMKGVYI | IAV | MS | 3 | 0.99 | 590 | 0.50 |  | 1/10 |
| **PA_649-658_** | LYASPQLEGF | IAV | MS/iedb | 1 | 0.99 | 59 | 0.81 | 5/10 | 0/10 |
| PB1_2-10_ | DVNPTLLFL | IAV | MS | 5 | 0.98 | 22015 | 0.29 |  | 4/10 |
| PB1_19-26_ | STTFPYTG | IAV | MS | 5 | 1 | 40250 | 0.09 |  | 0/10 |
| PB1_216-222_ | SYLIRAL | IAV | MS | 5 | 0.19 |  |  |  | 0/10 |
| PB1_216-224_ | SYLIRALTL | IAV | MS | 1 | 0.19 | 40 | 0.89 | 2/5 | 10/10 |
| PB1_430-438_ | RYTKTTYWW | IAV | MS | 1 | 0.15 | 26 | 0.98 | 2/5 | 0/10 |
| PB1_482-490_ | SYINRTGTF | IAV | MS | 1 | 0.16 | 23 | 1.03 | 0/5 | 0/10 |
| PB1_482-492_ | SYINRTGTFEF | IAV | MS | 1 | 0.14 | 20 | 1.07 | 0/5 | 0/10 |
| **PB1_496-505_** | FYRYGFVANF | IAV | iedb | 2 | 0.99 | 62 | 0.90 | 9/10 | 2/10 |
| **PB1_498-505_** | RYGFVANF | IAV | MS/iedb | 1 | 0.99 | 40 | 1.05 | 9/10 | 10/10 |
| PB1_500-505_ | GFVANF | IAV | MS | 5 | 0.99 |  |  |  | 0/10 |
| PB1-1_746-755_ | QYKGKLCL | IAV | MS | 4 |  | 18075 | 0.53 |  | 0/10 |
| PB1+2_8-16_ | EWMSIRPYF | IAV | MS | 2 |  | 88 | 0.64 | 1/5 | 0/10 |
| PB1+3_24-32_ | CYKHNFPLY | IAV | MS | 3 |  | 730 | 0.73 |  | 0/10 |
| PB1+3_682-689_ | QYGGGYGF | IAV | MS | 3 |  | 1134 | 0.81 |  | 1/10 |
| PB2_89-98_ | VMVSPLAVTW | IAV | MS | 3 | 0.91 | 2179 | 0.52 |  | 0/10 |
| PB2_110-119_ | HYPKIYKTYF | IAV | MS | 1 | 0.11 | 28 | 0.89 | 3/5 | 0/10 |
| PB2_112-119_ | PKIYKTYF | IAV | MS | 5 | 0.12 | 39253 | 0.32 |  | 0/10 |
| PB2_114-122_ | IYKTYFERV | IAV | MS | 2 | 0.01 | 157 | 0.74 | 0/5 | 0/10 |
| PB2_117-125_ | TYFERVERL | IAV | MS | 1 | 0.01 | 57 | 0.69 | 0/10 | 0/5 |
| PB2_204-212_ | AYMLERELV | IAV | MS | 2 | 0.99 | 194 | 0.49 | 0/5 | 0/10 |
| PB2_227-234_ | VYIEVLHL | IAV | MS | 2 | 0.59 | 361 | 0.87 | 0/5 | 1/10 |
| PB2_322-330_ | SFSFGGFTF | IAV | MS | 1 | 0.99 | 20 | 0.95 | 0/5 | 10/10 |
| PB2_437-446_ | HFQKDAKVLF | IAV | MS | 3 | 0.96 | 392 | 0.67 |  | 1/10 |
| PB2_463-471_ | ILPDMTPSI | IAV | MS | 3 | 0.01 | 1606 | 0.51 |  | 10/10 |
| PB2_549-555_ | TYQWIIR | IAV | MS | 5 | 0.97 |  |  |  | 0/10 |
| **PB2_549-557_** | TYQWIIRNW | IAV | MS | 2 | 0.96 | 62 | 0.86 | 4/8 | 10/10 |
| PB2_549-559_ | TYQWIIRNWET | IAV | MS | 4 | 0.17 | 8570 | 0.29 |  | 10/10 |
| PB2_552-559_ | WIIRNWET | IAV | MS | 5 | 0.17 | 36284 | 0.04 |  | 0/10 |
| PB2_591-599_ | QYSGFVRTL | IAV | MS | 2 | 0.44 | 214 | 0.61 | 1/5 | 1/10 |
| PB2_591-600_ | QYSGFVRTLF | IAV | MS | 1 | 0.44 | 20 | 0.85 | 0/5 | 0/5 |
| PB2_594-600_ | GFVRTLF | IAV | MS | 5 | 0.99 |  |  |  | 0/10 |
| PB2_703-710_ | RYGPALSI | IAV | MS | 2 | 0.98 | 389 | 0.88 | 1/5 | 4/10 |
| HA_176-184_ | TYPVLNVTM | IAV | MS | 2 | 0.01 | 351 | 0.70 | 1/5 | 0/10 |
| HA_248-259_ | IYWTIVKPGDVL | IAV | MS | 3 | 0.01 | 958 |  |  | 1/10 |
| HA_506-516_ | VYRDEALNNRF | IAV | MS | 3 | 0.40 | 556 | 0.68 |  | 0/10 |
| HA_507-516_ | YRDEALNNRF | IAV | MS | 5 | 0.41 | 20902 | 0.28 |  | 0/10 |
| HA_508-516_ | RDEALNNRF | IAV | MS | 5 | 0.41 | 28117 | 0.27 |  | 0/10 |
| M1_99-109_ | LYRKLKREITF | IAV | iedb | 3 | 0.48 | 409 | 0.70 |  | 0/10 |
| M1_108-117_ | TFHGAKEISL | IAV | iedb | 4 | 0.00 | 1274 | 0.39 |  | 0/10 |
| M1_239-248_ | AYQKRMGVQM | IAV | iedb | 4 | 0.66 | 3281 |  |  | 0/10 |
| NP_39-47_ | FYIQMCTEL | IAV | iedb | 2 | 0.99 | 61 | 0.63 | 1/10 | 6/10 |
| NP_138-148_ | IWHSNLNDATY | IAV | MS | 4 | 0.72 | 10945 | 0.54 |  | 0/10 |
| NP_257-265_ | TFLARSALI | IAV | MS | 2 | 0.12 | 218 | 0.65 | 1/5 | 0/10 |
| NP_296-304_ | YSLVGIDPF | IAV | MS | 3 | 0.9 | 1317 | 0.60 |  | 0/10 |
| NP_417-425_ | NLPFDRTTI | IAV | MS | 4 | 0.01 | 15720 | 0.34 |  | 2/10 |
| NP_419-429_ | PFDRTTIMAAF | IAV | iedb | 3 | 0.01 | 5016 | 0.51 |  | 0/10 |
| NP_456-464_ | VSFQGRGVF | IAV | MS | 4 | 0.14 | 7866 | 0.39 |  | 1/10 |
| NS2_98-106_ | TFMQALHLL | IAV | MS | 1 | 0.01 | 32 | 0.80 | 3/5 | 0/10 |

**Boldface** highlights immunodominant epitopes in human donors

Underline highlights immunogenic peptides in HHD-A24 mice

^1^Unique amino acid sequences of protein origin corresponding to the identified peptides from influenza A and B viruses were sourced from the NCBI database using full length sequences from human hosts isolated in Asia and/or Australia. Influenza A sequences were limited to H1N1 and H3N2 human isolate sequences. Sequence variants were identified using the *Identify short peptide in proteins* tool from the Influenza Research Database (www.fludb.org).

^2^ Analysed with NetpanMHC 4.0 (http://www.cbs.dtu.dk/services/NetMHCpan-4.0/)

^3^ Analysed with NetCTLpan 1.1 (http://www.cbs.dtu.dk/services/NetCTLpan/)

**Supplementary Table 3.** Influenza B peptides identified by immunopeptidomics

| Peptide | Sequence | Virus origin | Peptide pool | Conservation^1^ | Predicted binding affinity (nM)^2^ | Comb. CTL epitope score^3^ | Immunogenic in human donors (of tested) | Immunogenic in mice (of tested) |
| --- | --- | --- | --- | --- | --- | --- | --- | --- |
| PA_146-156_ | MIFSYNQDYSL | IBV | 11 | 0.99 | 2146 | 0.38 |  | 2/10 |
| PA_453-461_ | TVMMKYVLF | IBV | 11 | 0.99 | 122 | 0.58 |  | 0/14 |
| **PA_457-465_** | KYVLFHTSL | IBV | 10 | 0.96 | 220 | 0.72 | 8/14 | 0/14 |
| PB1_495-504_ | FYRDGFVSNF | IBV | 10 | 0.99 | 312 | 0.90 | 2/14 | 0/14 |
| **PB1_503-511_** | NFAMELPSF | IBV | 10 | 0.39 | 473 | 0.58 | 7/14 | 0/14 |
| PB2_103-113_ | TYGPIGDTEGF | IBV | 10 | 0.93 | 357 | 0.84 | 3/14 | 0/14 |
| PB2_245-253_ | IYHPGGNKL | IBV | 11 | 0.99 | 384 | 0.70 |  | 3/14 |
| PB2_405-413_ | VFSQDTRMF | IBV | 11 | 0.99 | 343 | 0.60 |  | 0/14 |
| PB2_433-441_ | MYQLQRYFL | IBV | 10 | 1 | 219 | 0.84 | 6/14 | 0/14 |
| PB2_439-448_ | YFLNRSNDLF | IBV | 10 | 0.65 | 63 | 0.87 | 5/14 | 0/14 |
| **PB2_550-558_** | TYQWVLKNL | IBV | 10 | 0.85 | 335 | 0.80 | 11/14 | 9/14 |
| HA_219-228_ | LYGDSKPQKF | IBV | 10 | 0.34 | 111 | 0.82 | 4/14 | 0/14 |
| HA_238-246_ | HYVSQIGGF | IBV | 11 | 0.68 | 728 | 0.62 |  | 0/14 |
| HA_338-346_ | IWVKTPLKL | IBV | 10 | 0.98 | 247 | 0.79 | 1/14 | 3/14 |
| HA_339-346_ | WVKTPLKL | IBV | 11 | 0.98 | 15986 | 0.30 |  | 1/10 |
| HA_370-380_ | GFLEGGWEGMI | IBV | 11 | 0.99 | 18807 | 0.44 |  | 1/9 |
| **HA_552-560_** | YYSTAASSL | IBV | 10 | 0.99 | 73 | 0.68 | 9/14 | 0/14 |
| HA_571-579_ | VYMVSRDNV | IBV | 11 | 0.89 | 2295 | 0.37 |  | 0/14 |
| HA+3_276-284_ | NYLSKRYFI | IBV | 10 | n.d. | 73 | 0.75 | 4/14 | 0/14 |
| M1_131-140_ | MYLNPGNYSM | IBV | 10 | 0.80 | 642 | 0.72 | 7/14 | 0/14 |
| NA_1-12_ | MLPSTIQTLTLF | IBV | 11 | 0.98 | 3376 | n.d. |  | 1/14 |
| NA_32-39_ | LYSDILLK | IBV | 11 | 0.91 | 24963 | 0.35 |  | 0/10 |
| **NA_32-40_** | LYSDILLKF | IBV | 10 | 0.91 | 44 | 1.04 | 9/14 | 14/14 |
| NA_209-217_ | AYTDTYHSY | IBV | 11 | 0.99 | 4132 | 0.62 |  | 0/13 |
| NA_213-221_ | TYHSYANNI | IBV | 10 | 0.25 | 65 | 0.69 | 6/14 | 11/14 |
| NA_402-411_ | SMEEPGWYSF | IBV | 11 | 0.63 | 943 | 0.60 |  | 1/14 |
| NA_458-466_ | TVTGVNMAL | IBV | 11 | 0.10 | 23313 | 0.31 |  | 0/10 |
| NA+3_480-489_ | PFVPILFEQL | IBV | 11 |  | 2296 | 0.59 |  | 0/14 |
| NP_79-89_ | VYNMVVKLGEF | IBV | 11 | 1 | 954 | 0.77 |  | 0/14 |
| **NP_164-173_** | IYFSPIRVTF | IBV | 10 | 0.17 | 25 | 1.00 | 11/14 | 14/14 |
| **NP_165-173_** | YFSPIRVTF | IBV | 10 | 0.17 | 65 | 1.05 | 12/14 | 14/14 |
| NP_181-191_ | MYKTTMGSDGF | IBV | 11 | 0.97 | 2637 | 0.68 |  | 0/12 |
| NP_218-228_ | VGLDPSLISTF | IBV | 11 | 1 | 8661 | 0.51 |  | 0/10 |
| NP_338-345_ | IYAKIPQL | IBV | 11 | 1 | 1015 | 0.90 |  | 0/12 |
| NP_338-347_ | IYAKIPQLGF | IBV | 10 | 1 | 9 | 1.01 | 7/14 | 4/14 |
| NP_392-400_ | AAYEDLRVL | IBV | 11 | 0.99 | 36058 | 0.29 |  | 13/14 |
| NP_399-408_ | VLSALTGTEF | IBV | 11 | 0.97 | 3729 | 0.45 |  | 0/12 |
| NS1_78-90_ | KAIGVKMMKVLLF | IBV | 11 | 0.95 | 6046 | n.d. |  | 0/12 |
| NS1_211-218_ | AYDQSGRL | IBV | 11 | 0.89 | 25406 | 0.38 |  | 0/10 |
| NS1_211-219_ | AYDQSGRLV | IBV | 11 | 0.89 | 17951 | 0.29 |  | 0/10 |
| NS2_28-37_ | VLMKDIQSQF | IBV | 11 | 0.89 | 1134 | 0.62 |  | 0/12 |

**Boldface** highlights immunodominant epitopes in human donors

Underline highlights immunogenic peptides in HHD-A24 mice

^1^Unique amino acid sequences of protein origins corresponding to the identified peptides from influenza A and B viruses were sourced from the NCBI database using full length sequences from human hosts isolated in Asia and/or Australia. Influenza A sequences were limited to H1N1 and H3N2 human isolate sequences. Sequence variants were identified using the *Identify short peptide in proteins* tool from the Influenza Research Database (www.fludb.org).

^2^ Analysed with NetpanMHC 4.0 (http://www.cbs.dtu.dk/services/NetMHCpan-4.0/)

^3^ Analysed with NetCTLpan 1.1 (http://www.cbs.dtu.dk/services/NetCTLpan/)

**Supplementary Table 4.** Screened IAV and IBV peptide variants

| Peptide | Sequence | Origin | Conservation^1^ |
| --- | --- | --- | --- |
| PB1_216-224_v1 | SYLIRALTL | IAV | 0.19 |
| PB1_216-224_v2 | GYLIRALTL | IAV | 0.79 |
| PB1_430-438_v1 | RYTKTTYWW | IAV | 0.15 |
| PB1_430-438_v2 | KYTKTIYWW | IAV | 0.28 |
| PB1_430-438_v3 | KYTKTTYWW | IAV | 0.55 |
| PB1_482-490_v1 | SYINRTGTF | IAV | 0.16 |
| PB1_482-490_v2 | SYINKTGTF | IAV | 0.83 |
| PB1_482-492_v1 | SYINRTGTFEF | IAV | 0.14 |
| PB1_482-492_v2 | SYINKTGTFEF | IAV | 0.83 |
| PB2_110-119_v1 | HYPKIYKTYF | IAV | 0.11 |
| PB2_110-119_v1 | HYPKVYKTYF | IAV | 0.87 |
| PB2_114-122_v1 | IYKTYFERV | IAV | 0.01 |
| PB2_114-122_v2 | VYKTYFEKV | IAV | 0.47 |
| PB2_114-122_v3 | IYKTYFEKV | IAV | 0.11 |
| PB2_114-122_v4 | VYKTYFDKV | IAV | 0.40 |
| PB2_117-125_v1 | TYFERVERL | IAV | 0.01 |
| PB2_117-125_v2 | TYFEKVERL | IAV | 0.58 |
| PB2_117-125_v3 | TYFDKVERL | IAV | 0.40 |
| PB2_227-234_v1 | VYIEVLHL | IAV | 0.59 |
| PB2_227-234_v2 | IYIEVLHL | IAV | 0.39 |
| PB2_227-234_v3 | MYIEVLHL | IAV | 0.01 |
| PB2_463-471_v1 | ILPDMTPSI | IAV | 0.01 |
| PB2_463-471_v2 | ILPDMTPST | IAV | 0.57 |
| PB2_463-471_v3 | VLPDMTPST | IAV | 0.40 |
| PB2_591-599_v1 | QYSGFVRTL | IAV | 0.44 |
| PB2_591-599_v2 | RYSGFVRTL | IAV | 0.46 |
| PB2_591-600_v1 | QYSGFVRTLF | IAV | 0.44 |
| PB2_591-600_v2 | RYSGFVRTLF | IAV | 0.46 |
| M1_99-109_v1 | LYRKLKREITF | IAV | 0.48 |
| M1_99-109_v2 | LYKKLKREITF | IAV | 0.49 |
| HA_176-184_v1 | TYPVLNVTM | IAV | 0.01 |
| HA_176-184_v2 | KYPALNVTM | IAV | 0.34 |
| HA_176-184_v3 | TYPALNVTV | IAV | 0.05 |
| HA_176-184_v4 | KYPVLNVTM | IAV | 0.01 |
| NP_257-265_v1 | TFLARSALI | IAV | 0.12 |
| NP_257-265_v2 | IFLARSALI | IAV | 0.80 |
| NP_257-265_v3 | IFSARSALI | IAV | 0.07 |
| NS2_98-106_v1 | TFMQALHLL | IAV | 0.01 |
| NS2_98-106_v2 | TFMQALQLL | IAV | 0.96 |
| NP_164-173_v1 | IYFSPIRVTF | IBV | 0.17 |
| NP_164-173_v2 | IYFSPIRITF | IBV | 0.79 |
| NP_165-173_v1 | YFSPIRVTF | IBV | 0.17 |
| NP_165-173_v2 | YFSPIRITF | IBV | 0.79 |
| NA_32-40_v1 | LYSDILLKF | IBV | 0.91 |
| NA_32-40_v2 | LYSDVLLKF | IBV | 0.01 |
| NA_213-221_v1 | TYHSYANNI | IBV | 0.25 |
| NA_213-221_v2 | TYHSYAKNI | IBV | 0.41 |
| NA_213-221_v3 | THYSYANKI | IBV | 0.25 |
| NA_213-221_v4 | THYSYAHNI | IBV | 0.01 |
| PB2_550-558_v1 | TYQWVLKNL | IBV | 0.85 |
| PB2_550-558_v2 | TYQWVMKNL | IBV | 0.14 |
| PB2_439-448_v1 | YFLNRSNDLF | IBV | 0.65 |
| PB2_439-448_v2 | YFLSRSNDLF | IBV | 0.34 |
| PB1_503-511_v1 | NFAMELPSF | IBV | 0.39 |
| PB1_503-511_v2 | NFAMEIPSF | IBV | 0.61 |
| M1_131-140_v1 | MYLNPGNYSM | IBV | 0.80 |
| M1_131-140_v2 | MYLNRGNYSM | IBV | 0.02 |
| M1_131-140_v3 | MYLNPENYSM | IBV | 0.02 |
| HA_219-228_v1 | LYGDSKPQKF | IBV | 0.34 |
| HA_219-228_v2 | LYGDSNPQKF | IBV | 0.61 |

^1^Unique amino acid sequences of protein origins of the identified peptides of influenza A and B viruses were sourced from the NCBI database using full length sequences from human hosts isolated in Asia and/or Australia. Influenza A sequences were limited to H1N1 and H3N2 human isolate sequences. Sequence variants were identified using the *Identify short peptide in proteins* tool from the Influenza Research Database (www.fludb.org).

**Supplementary Table 5.** Data Collection, Refinement Statistics and Thermal stability of peptide-HLA complexes

| **Data Collection Statistics** | **HLA-A*24:02-PB2_549-559_** | **HLA-A*24:02-**  **PB2_549-557_** | **HLA-A*24:02-PB2_549-557B_** | **HLA-A*24:02-NP_165-173_** | **HLA-A*24:02-NP_164-173_** |
| --- | --- | --- | --- | --- | --- |
| Space group | **I2** | **P6_5_22** | **P2_1_** | **P2_1_2_1_2_1_** | **C222_1_** |
| Cell Dimensions (a,b,c) (Å) | 89.68, 43.65, 236.49, β=95.56° | 87.29, 87.29, 312.46 | 46.07, 123, 86.59  β=103.4° | 62.95, 75.55, 89.91 | 79.92, 120.25, 187.80 |
| Resolution (Å) | 39.23 – 2.95  (3.13 – 2.95) | 48.16 – 2.90  (3.08 – 2.90) | 44.81 – 2.16 (2.23 – 2.16) | 34.91 – 1.51  (1.54 – 1.51) | 46.95 – 2.75  (2.90 – 2.75) |
| Total number of observations | 73078 (11941) | 159459 (25130) | 188348 (15744) | 625371 (30507) | 158168 (24193) |
| Number of unique observations | 19624 (3149) | 16623 (2577) | 49524 (4188) | 64117 (3233) | 23763 (3421) |
| Multiplicity | 3.7 (3.8) | 9.6 (9.8) | 3.8 (3.8) | 9.8 (9.4) | 6.7 (7.1) |
| Data completeness (%) | 99.6 (99.4) | 99.9 (99.9) | 99.2 (96.9) | 94.0 (96.2) | 99.4 (99.3) |
| I/σ_I_ | 12.3 (2.0) | 16.4 (2.1) | 14.4 (2.6) | 28.4 (4.4) | 7.8 (1.9) |
| R_pim_^a^ (%) | 6.7 (40.4) | 2.9 (31.1) | 4.6 (31.9) | 1.4 (14.9) | 7.5 (40.3) |
| **Refinement Statistics** |  |  |  |  |  |
| Non-hydrogen atoms | 6405 | 3177 | 6898 | 3914 | 6387 |
| Protein | 6328 | 3159 | 6309 | 3320 | 6295 |
| Water | 77 | 14 | 587 | 594 | 91 |
| *R_factor_*^b^ (%) | 20.3 | 24.1 | 17.8 | 18.2 | 18.9 |
| *R_free_*^b^ (%) | 27.6 | 27.4 | 22.9 | 20.6 | 26.1 |
| Rms deviations from ideality |  |  |  |  |  |
| Bond lengths (Å) | 0.010 | 0.007 | 0.010 | 0.006 | 0.010 |
| Bond angles (°) | 1.16 | 0.96 | 1.08 | 0.87 | 1.16 |
| Ramachandran plot (%) |  |  |  |  |  |
| Favoured region | 91.0 | 93.0 | 99.3 | 98.0 | 94.0 |
| Allowed region | 8.0 | 6.0 | 0.7 | 2.0 | 5.0 |
| Disallowed region | 1.0 | 1.0 | 0.0 | 0.0 | 0.0 |
| **Tm (℃)** | 57.1 ± 1.3 | 61.9 ± 1.3 | 57.1 ± 1.2 | 57.5 ± 0.3 | 64.0 ± 2.2 |

^a^R_p.i.m_ = Σ_hkl_ [1/(N-1)]^1/2^ Σ_i_ | I_hkl, i_ - <I_hkl_> | / Σ_hkl_ <I_hkl_>, ^b^ R_factor_ = Σ_hkl_ | | F_o_ | - | F_c_ | | / Σ_hkl_ | F_o_ | for all data except ≈ 5% which were used for R_free_ calculation.

**Supplementary Table 6.** Staining panels

| Panel 1 (Mouse ICS) | | | |
| --- | --- | --- | --- |
| **Antibody** | **Flourochrome** | **Vendor (cat. No)** | **Dilution** |
| Live/Dead | NIR | Invitrogen (L34976) | 1:800 |
| CD8 | PerCP-Cy 5.5 | BD Pharmingen (551162) | 1:350 |
| CD4 | PE-Cy7 | eBioscience (25-0041-82) | 1:200 |
| IL-2 | PE | BD Pharmingen (554420) | 1:200 |
| TNF | APC | BD Pharmingen (554428) | 1:200 |
| IFNγ | FITC | BioLegend (505806) | 1:200 |

| Panel 2 (Human ICS) | | | |
| --- | --- | --- | --- |
| **Antibody** | **Flourochrome** | **Vendor (cat. No)** | **Dilution** |
| Live/Dead | NIR | Invitrogen (L34976) | 1:800 |
| CD3 | PE-Cy7 | BD Pharmingen (563423) | 1:50 |
| CD4 | PE | BD Pharmingen (555347) | 1:50 |
| CD8 | PerCP-Cy5.5 | BD Pharmingen (565310) | 1:100 |
| CD107a | AF488 | Invitrogen (53107942) | 1:200 |
| IFNγ | BV421 | BD Horizon (560371) | 1:100 |
| TNF | AF700 | BD Pharmingen (557996) | 1:50 |
| MIP-1β | APC | BD Pharmingen (560686) | 1:40 |

| Panel 3 (PBMC TAME) | | | |
| --- | --- | --- | --- |
| **Antibody** | **Flourochrome** | **Vendor (cat. No)** | **Dilution** |
| CD71 | BV421 | BD Horizon (562995) | 1:50 |
| CD3 | BV510 | BioLegend (317332) | 1:200 |
| HLA-DR | BV605 | BioLegend (307640) | 1:100 |
| CD4 | BV650 | BD Horizon (563875) | 1:200 |
| CD27 | BV711 | BD Horizon (563167) | 1:200 |
| CD38 | BV786 | BD Horizon (563964) | 1:100 |
| Tetramer 1 | APC | In house |  |
| CCR7 | AF700 | BD Pharmingen (561143) | 1:50 |
| CD14 | APC-H7 | BD Pharmingen (561143) | 1:100 |
| CD19 | APC-H7 | BD Pharmingen (560177) | 1:100 |
| Live/Dead | NIR | Invitrogen (L34976) | 1:800 |
| CD45RA | FITC | BD Pharmingen (555488) | 1:200 |
| CD8 | PerCp-Cy5.5 | BD Pharmingen (565310) | 1:50 |
| Tetramer 2 | PE | In house |  |
| CD95 | PE-CF594 | BD Horizon (561272) | 1:100 |
| PD1 | PE-Cy7 | BD Pharmingen (561272) | 1:50 |

| Panel 4 (Tissue TAME) | | | |
| --- | --- | --- | --- |
| **Antibody** | **Flourochrome** | **Vendor (cat. No)** | **Dilution** |
| CD69 | BV421 | BioLegend (310930) | 1:100 |
| CD3 | BV510 | BioLegend (317332) | 1:200 |
| HLA-DR | BV605 | BioLegend (307640) | 1:100 |
| CD4 | BV650 | BD Horizon (563875) | 1:200 |
| CD27 | BV711 | BD Horizon (563167) | 1:200 |
| CD38 | BV786 | BD Horizon (563964) | 1:100 |
| Tetramer 1 | APC | In house |  |
| CCR7 | AF700 | BD Pharmingen (561143) | 1:50 |
| CD14 | APC-Cy7 | BD Pharmingen (561143) | 1:100 |
| CD19 | APC-Cy7 | BD Pharmingen (560177) | 1:100 |
| Live/Dead | NIR | Invitrogen (L34976) | 1:800 |
| CD103 | FITC | BioLegend (350203) | 1:100 |
| CD8 | PerCp-Cy5.5 | BD Pharmingen (565310) | 1:50 |
| Tetramer 2 | PE | In house |  |
| CD95 | PE-CF594 | BD Horizon (561272) | 1:100 |
| CD45RO | PE-Cy7 | ThermoFisher (25-0457-41) | 1:100 |
